# Identification of novel biomarkers for pyridoxine-dependent epilepsy using untargeted metabolomics and infrared ion spectroscopy - biochemical insights and clinical implications

**DOI:** 10.1101/2021.01.22.20248925

**Authors:** Udo F.H. Engelke, Rianne E. van Outersterp, Jona Merx, Fred A.M.G. van Geenen, Arno van Rooij, Giel Berden, Marleen C.D.G. Huigen, Leo A.J. Kluijtmans, Tessa M.A. Peters, Hilal H. Al-Shekaili, Blair R. Leavitt, Erik de Vrieze, Sanne Broekman, Erwin van Wijk, Laura A. Tseng, Purva Kulkarni, Floris P.J.T. Rutjes, Jasmin Mecinović, Eduard A. Struys, Laura A. Jansen, Sidney M. Gospe, Saadet Mercimek-Andrews, Keith Hyland, Michèl A.A.P. Willemsen, Levinus A. Bok, Clara D.M. van Karnebeek, Ron A. Wevers, Thomas J. Boltje, Jos Oomens, Jonathan Martens, Karlien L.M. Coene

**Author notes:** First three and last two authors contributed equally (Udo F.H. Engelke, Rianne E. van Outersterp, Jona Merx, Jonathan Martens, Karlien L.M. Coene).

## Abstract

Pyridoxine-dependent epilepsy (PDE-ALDH7A1), also known as antiquitin deficiency, is an inborn error of lysine metabolism that presents with refractory epilepsy in newborns. Bi-allelic *ALDH7A1* variants lead to deficiency of α-aminoadipic semialdehyde dehydrogenase, resulting in accumulation of piperideine-6-carboxylate (P6C), and secondary deficiency of the important co-factor pyridoxal-5’-phosphate (PLP, active vitamin B6) through its complexation with P6C. Vitamin B6 supplementation resolves epilepsy in patients, but despite this treatment, intellectual disability may occur. Early diagnosis and treatment, preferably based on newborn screening, potentially optimize long-term clinical outcome. However, the currently known diagnostic PDE-ALDH7A1 biomarkers are incompatible with newborn screening procedures. Using a combination of the innovative analytical methods untargeted metabolomics and infrared ion spectroscopy, we have been able to discover novel biomarkers for PDE-ALDH7A1: 2S,6S-and 2S,6R-oxopropylpiperidine-2-carboxylic acid (2-OPP) and 6-oxopiperidine-2-carboxylic acid (6-oxoPIP). We demonstrate the applicability of 2-OPP as a PDE-ALDH7A1 biomarker in newborn screening. Additionally, we show that 2-OPP accumulates in brain tissue of patients and *Aldh7a1* knock-out mice, and induces epilepsy-like behavior in a zebrafish model system. We speculate that 2-OPP may contribute to ongoing neurotoxicity, also in treated PDE-ALDH7A1 patients. As 2-OPP formation appears to increase upon ketosis, we emphasize the importance of avoiding catabolism in PDE-ALDH7A1 patients.

## Introduction

Pyridoxine-dependent epilepsy due to bi-allelic *ALDH7A1* variants (OMIM 266100, PDE-ALDH7A1) is an inborn error of metabolism (IEM) that presents with refractory seizures in early infancy. These seizures are typically unresponsive to anti-convulsant medications, but can be controlled with supplementation of pyridoxine (vitamin B6). *ALDH7A1* encodes the enzyme α-aminoadipic semialdehyde (α-AASA) dehydrogenase, which is also known as antiquitin. Deficiency of this enzyme disrupts lysine catabolism at the level of the conversion of α-AASA to α-aminoadipic acid (Figure 1), leading to accumulation of α-AASA and pipecolic acid in body fluids (1, 2). α-AASA is found in equilibrium with its alternative molecular form Δ1-piperideine-6-carboxilic acid (P6C), which is formed by spontaneous cyclization. Subsequent condensation of accumulating P6C with pyridoxal 5’-phosphate (PLP) results in secondary deficiency of active vitamin B6, which is thought to be the main pathophysiological mechanism in PDE-ALDH7A1 (3). By supplementing patients with vitamin B6, this secondary deficiency can be corrected. Vitamin B6 supplementation is therefore a cornerstone of seizure treatment in PDE-ALDH7A1, although some patients may require add-on anti-convulsant medication for optimal seizure control (4). Recent studies have shown that a lysine-restricted diet can also be beneficial for ensuring optimal developmental outcome in these patients, but only if such a diet is started in the first year of life (5-8). As well, the addition of arginine supplementation to this treatment regimen, referred to as triple therapy, has been shown to improve neurodevelopmental outcome when started early in life (7, 8). Arginine competes with lysine for transport across the blood-brain barrier and therefore lowers the flux through lysine catabolism in the brain. At the biochemical level, triple therapy proved to lower levels of P6C, α-AASA and pipecolic acid in plasma and cerebrospinal fluid (CSF) of treated PDE-ALDH7A1 patients (7, 8).

**Figure 1.**
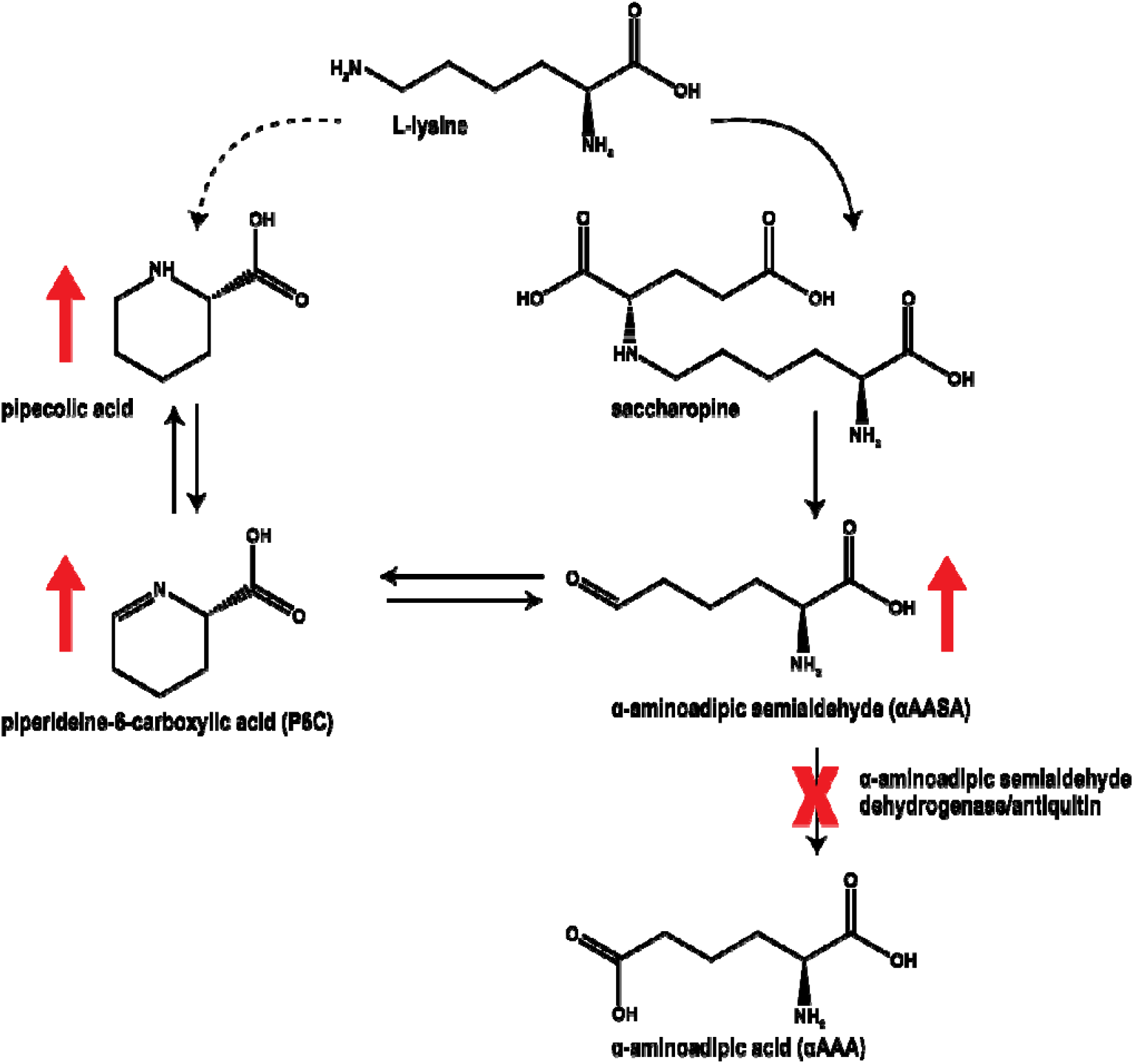
Overview of the L-lysine metabolic pathway involved in pyridoxine-dependent epilepsy. Diminished function of the enzyme α-aminoadipic semialdehyde dehydrogenase (antiquitin) coded by the *ALDH7A1* gene (indicated by red cross) leads to accumulation (indicated by red upward arrows) of α-aminoadipic semialdehyde (α-AASA), its cyclic counterpart piperideine-6-carboxylic acid (P6C) and pipecolic acid.

In order to optimize clinical outcome, it is crucial that the diagnosis of PDE-ALDH7A1 is made as early as possible in life. The combination of readily available disease-modifying therapeutic options, the potential risk of additional brain damage upon delayed diagnosis, along with a reasonably high incidence of this rare disease (estimated 1:64000 conceptions (9)), makes PDE-ALDH7A1 an eligible condition for inclusion in newborn screening programs. However, this is presently not implemented as optimal biomarkers for dried blood spot (DBS) analysis in newborn screening are not available. The current tools for rapid diagnostic screening for PDE-ALDH7A1 encompass analysis of α-AASA and P6C in urine (10). Measurement of α-AASA and P6C in plasma and DBS has proven to be challenging because of their limited stability at room temperature (11). Analytical methods have been published that attempt to circumvent this, however, these are incompatible with commonly used newborn screening workflows, which make use of direct infusion MS methods without LC separation in order to avoid time-consuming derivatization steps to enable high-throughput (12-14). Therefore, there is a clear clinical need for other biomarkers to be able to include PDE-ALDH7A1 in newborn screening. Additionally, clinical observations in PDE-ALDH7A1 patients suggest that our current understanding of disease pathophysiology is likely incomplete. Intellectual disability is present in many patients despite seizure control with pyridoxine, and in a subset of patients, progressive brain MRI abnormalities are observed with enlargement of ventricles and progressive white matter and corpus callosum abnormalities (15-19). In episodes of intercurrent febrile illness, break-through epileptic seizures can occur despite ongoing vitamin B6 treatment. In fact, according to data from the PDE registry (http://www.pdeonline.org/, (5)), break-through seizures manifest in almost 40% of the 116 patients enlisted in the registry. Also, a substantial percentage of patients still requires add-on classical anti-convulsant drugs (18% based on PDE registry data) (4). The molecular mechanisms underlying all these clinical observations are unknown, but they are suggestive of biochemical disease sequelae that go beyond our current understanding of disrupted lysine metabolism in PDE-ALDH7A1.

To address both the need for novel diagnostic biomarkers for PDE-ALDH7A1, as well as the need for deeper insights in the biochemical effects of α-AASA dehydrogenase deficiency, unbiased screening of the metabolic profile of patient body fluids is a crucial step. This can now be achieved through an innovative high-resolution mass spectrometry (MS) based technique referred to as ‘untargeted metabolomics’. In our laboratory, we have established an untargeted metabolomics method for broad diagnostic screening of IEMs, coined Next Generation Metabolic Screening (NGMS) (20), which we have now applied in search of novel biomarkers for PDE-ALDH7A1. We have previously shown the potential of such an approach in the identification of novel IEM biomarkers and increasing pathophysiological insights in disease (21, 22). However, the structural elucidation of unknown features/novel metabolites from untargeted metabolomics data remains a significant challenge. To overcome this challenge, we used a unique combination of NGMS and infrared ion spectroscopy (IRIS) for orthogonal structural identification (23-28). Using this innovative methodology, we were able to identify novel, discriminative biomarkers for PDE-ALDH7A1, which can be measured in urine, plasma, and CSF, as well as in DBS. Our findings further pave the way for inclusion of PDE-ALDH7A1 in newborn screening, increase our insights in underlying disease mechanisms and may pose clinical implications for patient management.

## Results

### Untargeted metabolomics/NGMS identifies novel biomarkers in body fluids of PDE-ALDH7A1 patients

We performed untargeted metabolomics/NGMS in 11 plasma samples from 7 PDE-ALDH7A1 patients (3 female and 4 male patients, mean age at sampling 14 years). Upon selection of features that were significantly increased in all patient plasma samples compared to non-IEM controls, we could readily identify features corresponding to the known biomarkers α-ASAA, P6C and pipecolic acid. Additionally, two isobaric features with a mass over charge ratio (m/z) of 186.1123 and retention times (RT) of 2.33 min (biomarker A, fold change 200x compared to controls) and 2.55 min (biomarker B, fold change 145x compared to controls), as well as a third feature with a m/z of 144.0652 and RT of 3.05 min, (biomarker C, fold change 45x compared to controls) were identified as significantly increased in all patient samples. Subsequently, in CSF and urine samples of PDE-ALDH7A1 patients, we were also able to detect these unknown features in significantly increased levels. An overview of these findings is depicted in Table 1. Metabolite annotation of these features based on their accurate mass using the Human Metabolome Database (HMDB) only rendered ecgonine (HMDB0006548) and pseudoecgonine (HMDB0006348) as a match for biomarkers A and B. These are tropane alkaloids related to cocaine, so we considered this identification to be very unlikely. For biomarker C, HMDB indicated multiple possible matches to the exact mass, however, one of these matches, 6-oxopiperidine-2-carboxylic acid (6-oxoPIP, HMDB 0061705), stood out as a likely candidate because of its chemical similarity to P6C.

**Table 1.** Untargeted metabolomics/NGMS results for biomarkers in PDE-ALDH7A1 patient body fluids. All six features shown in Table 1 were found to be significantly increased in PDE-ALDH7A1 patient plasma, urine and CSF compared to controls. The mean fold change in intensity in plasma is indicated, and the adduct with the highest fold change is shown, either the [M+H] ^+^ adduct generated in the positive ionization mode, or the [M-H] adduct from the negative ionization mode. m/z: mass over charge ratio, RT (min): retention time in minutes.

| *Feature* | *m/z* | *RT (min)* | *Fold change* |
| --- | --- | --- | --- |
| *Biomarker A [M+H]<sup>+</sup>* | 186.1123 | 2.33 | 200 |
| *Biomarker B [M+H]<sup>+</sup>* | 186.1123 | 2.55 | 145 |
| *Biomarker C [M+H]<sup>+</sup>* | 144.0652 | 3.05 | 45 |
| *α-ASAA [M-H]<sup>-</sup>* | 144.0666 | 0.86 | 8 |
| *P6C [M+H]<sup>+</sup>* | 128.0706 | 0.86 | 9 |
| *Pipecolic acid [M+H]<sup>+</sup>* | 130.0861 | 1.04 | 3 |

For molecular identification of biomarkers A and B, we turned to a novel approach using IRIS, which our group has recently successfully applied for metabolite identification (23, 24, 27). In short, MS fragmentation of biomarkers A and B gave a first hint to their chemical composition, as an MS fragment corresponding to P6C could be detected with an m/z of 128.0706. Through the comparison of *in silico* generated IR spectra for putative chemical structures of biomarkers A and B to their experimental IR spectra as measured from PDE-ALDH7A1 patient plasma, we were able to further pinpoint their chemical structure without using reference standards. Based on this preliminary structural assignment using predicted IR spectra, we could focus on synthesis of only the predetermined compounds assigned to biomarkers A and B for use in final confirmation of their structures, and as analytical standards for the development of targeted MS-based diagnostics. Using this approach, we were able to identify biomarkers A and B as two diastereomers of 2-oxopropylpiperidine-2-carboxylic acid (2-OPP, 2S,6S and 2S,6R conformations, respectively) (Fig 2A-B). A detailed description of the IRIS strategy for molecular identification of the 2-OPP diastereomers will be described separately by van Outersterp *et al*. (manuscript in preparation). For biomarker C, a reference standard of 6-oxoPIP was commercially available, and by comparison of its IR spectrum to the IR spectrum measured for the signal observed in patient body fluids, we were able to confirm the identity of biomarker C as 6-oxoPIP (Fig 2C). During the preparation of this manuscript, 6-oxoPIP has been reported by two independent studies as a novel PDE-ALDH7A1 biomarker (29, 30). However, its suitability for detection in DBS for neonatal screening was not further evaluated.

**Figure 2.**
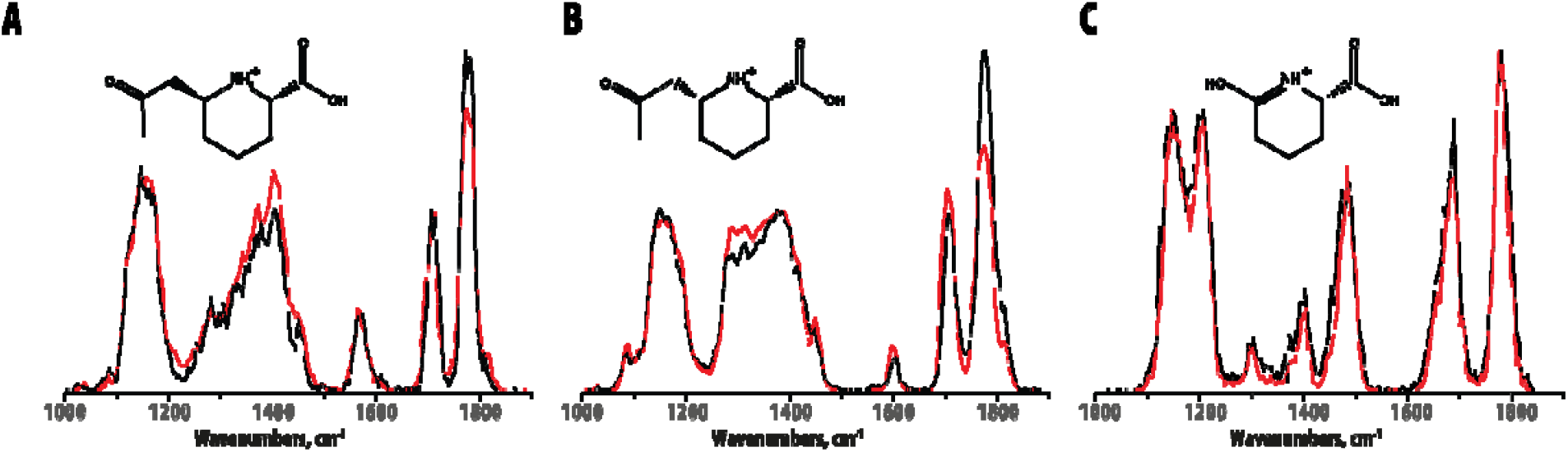
Molecular identification of unknown biomarkers in PDE-ALDH7A1 from untargeted metabolomics/NGMS data using IRIS. Matching overlay of the IR spectra of biomarker A (m/z 186.1123 and RT 2.33 min, panel A), biomarker B (m/z 186.1123 and RT 2.55 min, panel B) and biomarker C (m/z 144.0652 and RT 3.05, panel C) measured in PDE-ALDH7A1 patient plasma (black traces) and the IR spectra of the protonated 2S,6S-2OPP (panel A), 2S,6R-2-OPP (panel B) and 6-oxoPIP (panel C) ions measured from synthetic reference standards (red traces). Molecular structures of the ions are inlayed in each panel.

### Quantitative LC-MS measurements of 6-oxoPIP and 2-OPP in PDE-ALDH7A1 patient body fluids

To further examine the suitability of 2-OPP and 6-oxoPIP as possible biomarkers for PDE-ALDH7A1, we developed quantitative targeted assays for these markers, using LC-tandem MS for analysis in plasma, urine, and CSF. These methods are further described in the Methods section of this paper and were validated according to ISO:15189 criteria. In brief, we analyzed 11 available plasma samples, 8 urine samples and 9 CSF samples from a total of 15 PDE-ALDH7A1 patients (8 males, 7 females, mean age at sampling 13 years) who all had a genetically confirmed diagnosis. These patients were on different treatment regimens (or untreated) and we compared 6-oxoPIP and 2-OPP levels (the sum of the 2-OPP 2S,6S and 2S,6R diastereomers was used) to anonymized, non-PDE control samples, which were age-matched as much as possible (Figure 3). The quantitatively measured levels of 6-oxoPIP and 2-OPP in plasma showed satisfactory correlation to the corrected semi-quantitative intensities measured using NGMS as described above (Supplemental figure 1). In urine of PDE-ALDH7A1 patients, mean 6-oxoPIP levels were substantially higher than 2-OPP levels (mean 6-oxoPIP 117.4 μM/mmol creatinine versus 2-OPP 2.6 μM/mmol creatinine). This difference was also observed in plasma, though to a lesser extent (mean 6-oxoPIP 7.5 μM versus mean 2-OPP 3.1 μM). However, in CSF, the difference in concentrations was the other way around (mean 6-oxoPIP 0.5 μM versus mean 2-OPP 2.1 μM). Overall, we found no clear within-sample correlation between 6-oxoPIP and 2-OPP levels (Supplemental figure 2), however, a positive correlation of 6-oxoPIP and 2-OPP in urine was found with the known PDE-ALDH7A1 urinary biomarker α-AASA (Supplemental figure 3). In all measured body fluids, PDE-ALDH7A1 patient samples could be distinguished from controls based on increased concentrations of the novel biomarkers 2-OPP and 6-oxoPIP.

**Figure 3.**
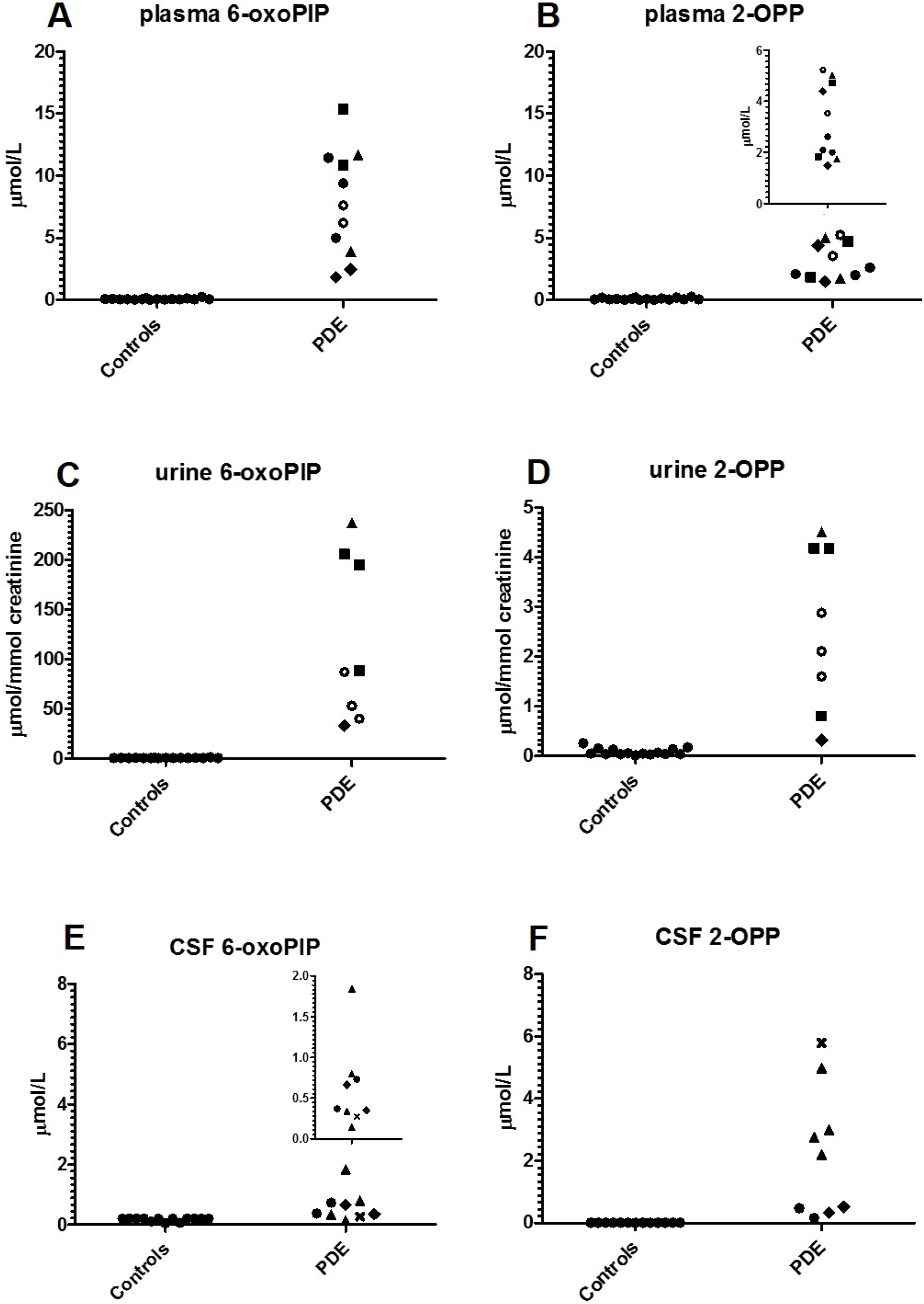
Absolute concentration of 6-oxoPIP and the sum of 2S,6S-2-OPP and 2S,6R-2-OPP in plasma **(A**,**B)**, urine **(C**,**D)** and CSF **(E**,**F)** of PDE-ALDH7A1 patients (PDE) compared to non-IEM controls (Controls), as measured by quantitative LC-MS/MS. Different treatment regimens in patients are coded as follows: open circles: untreated; filled squares: vitamin B6 supplementation; filled circles: vitamin B6 and arginine supplementation; filled triangles: vitamin B6 supplementation and lysine restriction; filled diamonds: vitamin B6 and arginine supplementation and lysine restriction; cross: therapy unknown. In B and E, the inserts show a close-up of the concentration range measured in patients.

### 2-OPP and 6-oxo-PIP are increased in PDE-ALDH7A1 patient brain tissue and in plasma and brain tissue *Aldh7a1* knock-out mice

To further evaluate biological relevance of the novel PDE biomarkers, we analyzed an available brain tissue extract sample, obtained post-mortem from a deceased PDE-ALDH7A1 patient, using our NGMS assay (Figure 4A-E). The clinical details of this deceased patient have been described previously (18, 31). In this brain tissue extract, we were able to confirm the presence of features corresponding to the known PDE markers P6C, α-AASA and pipecolic acid (Figure 4A-B, E). Additionally, the novel markers 2-OPP (2S,6S and 2S,6R diastereomers) and 6-oxoPIP could be observed in the PDE brain tissue extract (Figure 4C-D). Upon quantitative measurement, we determined that 2-OPP levels were ∼5 nmol/gram cortical tissue and 6-oxoPIP levels were ∼29 nmol/gram. In a previous study, a P6C concentration of 8.14 nmol/gram was reported in this brain specimen, and α-AASA was determined to be 13.02 nmol/gram (31). In brain extracts of four non-PDE controls, 2-OPP and 6-oxoPIP levels were below the detection limit of our quantitative assay. In a recently described *Aldh7a1* knock-out (KO) mouse model (32), we were also able to detect increased concentrations of 2-OPP and 6-oxoPIP in plasma (Fig 5A-B) and brain extracts (Figure 5E-F) of KO mice, supporting our findings from the human situation. The known PDE-ALDH7A1 markers P6C, α-AASA and pipecolic acid (Figure 5C-D,G) were also found to be increased in brain tissue of *Aldh7a1* KO mice, with pipecolic acid accumulating in remarkably high levels.

**Figure 4.**
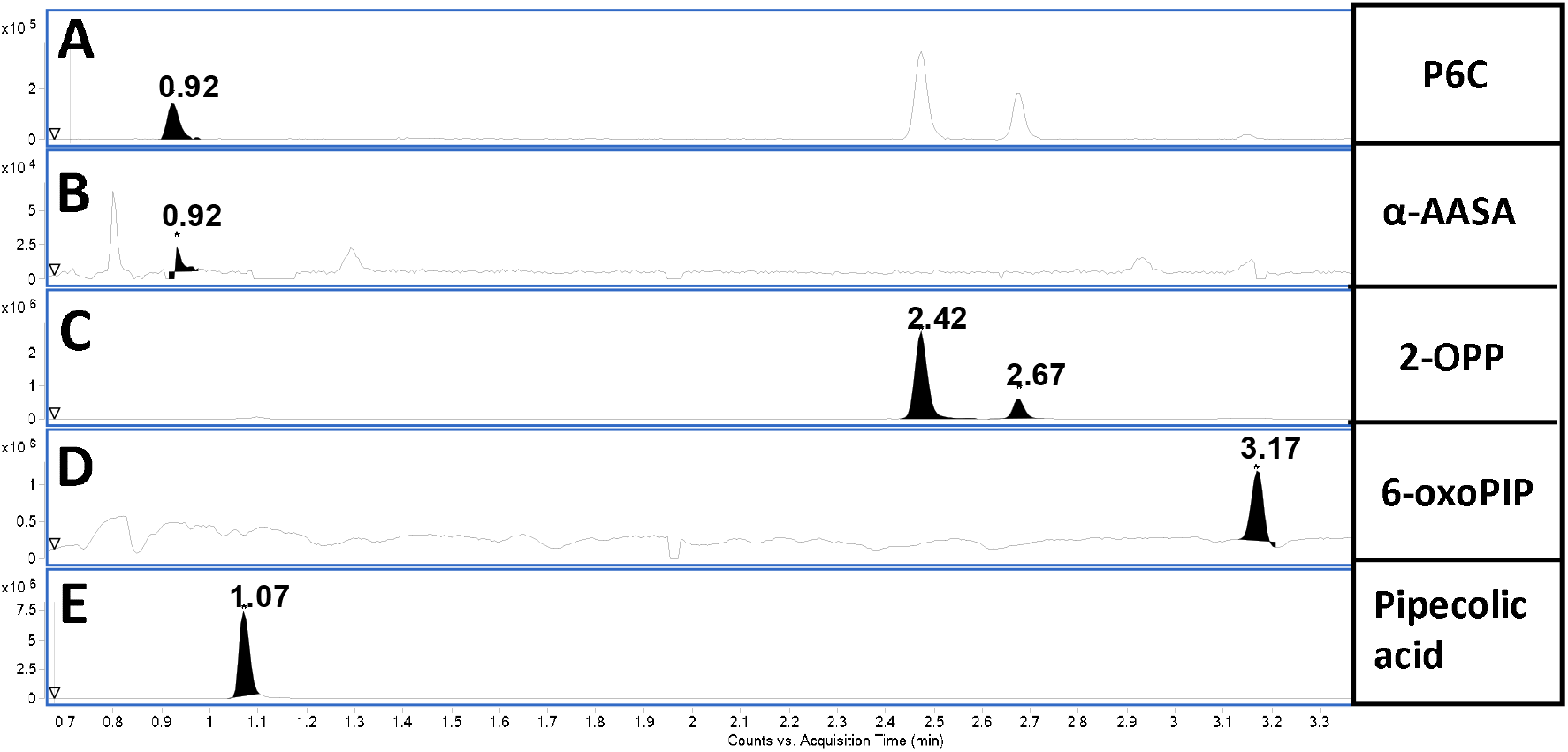
Extracted Ion Chromatograms (EIC) in brain tissue extract of a PDE-ALDH7A1 patient of **A**. P6C, m/z 128.0706, RT 0.92 min, **B**. α-AASA, m/z 146.0812, RT 0.92 min, **C**. 2S,6S-2-OPP and 2S,6R-2-OPP, respectively, m/z 186.1123, RT 2.42 and 2.67 min, **D**. 6-oxoPIP, m/z 144.0652, RT 3.17 min, and E. pipecolic acid, m/z 130.0863, RT 1.07 min. Y-axis represents relative NGMS intensity, X-axis represents RT (min). Note the fact that also an in-source P6C-fragment of both 2-OPP enantiomers is observed at RT 2.42 and 2.67 min (**A**).

**Figure 5.**
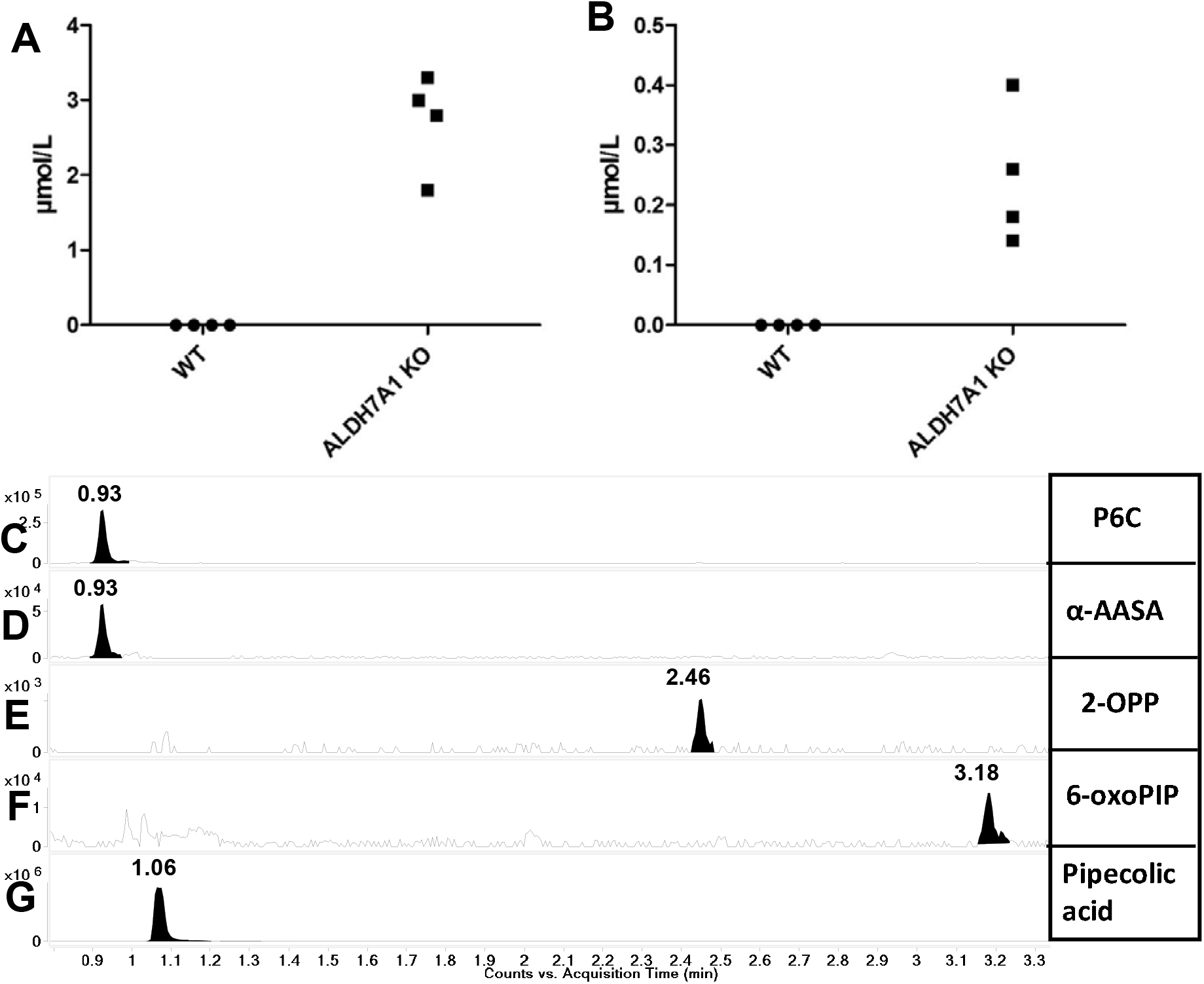
Plasma concentrations in *Aldh7A1* KO mice compared to wildtype (WT) mice of **A**. 6-oxoPIP, and **B**. the sum of 2S,6S-2-OPP and 2S,6R-2-OPP; Extracted Ion Chromatograms (EIC) in brain tissue extract of *Aldh7a1* KO mice of **C**. P6C, m/z 128.0706, RT 0.93 min, **D**. α-AASA, m/z 146.0812, RT 0.93 min, **E**. 2S,6S-2-OPP, m/z 186.1125, RT 2.46, **F**. 6-oxoPIP, m/z 144.0652, RT 3.18 min, and **G**. pipecolic acid, m/z 130.0863, RT 1.06 min. Y-axis represents relative NGMS intensity, X-axis represents RT (min).

### 2-OPP is formed by reaction of P6C with acetoacetate under physiological conditions

To gain further insight in the biochemical origin of 2-OPP as a biomarker for α-AASA dehydrogenase deficiency, we performed additional experiments to mimic physiological conditions for its formation. From the chemical structure of 2-OPP, as well as from the chemical synthesis of the 2-OPP model compound, we envisioned that 2-OPP could likely be formed in the body by the reaction of accumulating P6C with the ketone body acetoacetate (Figure 6H). To further substantiate this hypothesis, we incubated P6C with acetoacetate under physiological conditions in both water, phosphate buffered saline (PBS), human (non-IEM) control plasma and human control urine (Figure 6A-G). In all these incubation experiments we observed preferential formation of the 2S,6S-2-OPP enantiomer, however, in plasma there appeared to be comparable levels of both the 2S,6R- and 2S,6S-2-OPP enantiomers. Additionally, we incubated P6C with plasma and urine of a patient who was in ketotic state (based on increased concentration of acetyl-carnitine in plasma and 3-hydroxy-butyric acid in urine; acetoacetate itself was not measured but was assumed to be elevated due to ketosis), and found that 2-OPP was readily formed in both samples, with the ratio of isomers similar to that found in the incubation experiments in control urine and plasma as described above. Finally, we incubated P6C in control plasma without the addition of acetoacetate and found detectable but significantly lower levels of 2-OPP formed. We therefore hypothesize that, even in non-ketotic circumstances, low amounts of acetoacetate present in body fluids of PDE-ALDH7A1 patients can react with accumulating P6C to form micromolar levels of 2-OPP. The identity of the observed features as 2-OPP was confirmed by obtaining IR spectra of the two 2-OPP features and matching those to reference IR spectra (Supplemental Figure 4).

**Figure 6.**
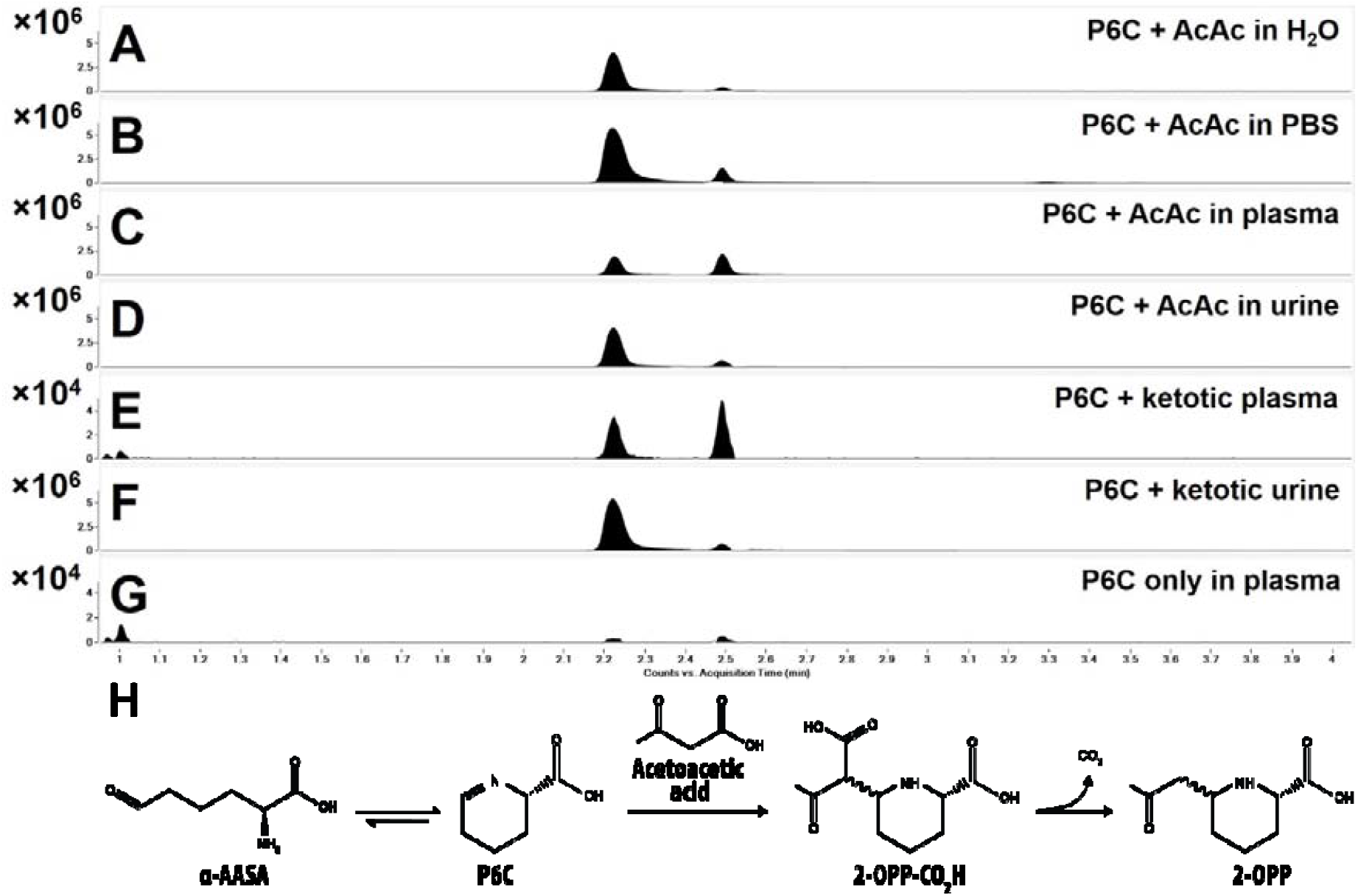
2-OPP is formed from P6C and acetoacetate (AcAc). Upon incubation of 1mM P6C with 1mM AcAc in different matrices, being H_2_O (**A**), PBS (**B**), control plasma (**C**) and control urine (**D**), 2-OPP formation was observed at elevated levels. Also upon incubation of 1mM P6C with ketotic plasma (**E**) and urine (**F**), elevated 2-OPP formation was observed. In the negative control incubation in plasma, including P6C but without the addition of AcAc, significantly lower 2-OPP signals were detected (**G**). Y-axis represents relative NGMS intensity (10^4^ or 10^6^ scale), X-axis represents RT (min). (**H**) The proposed chemical reaction of formation of 2-OPP from AcAc and P6C (P6C is formed in equilibrium from αAASA).

### 2-OPP shows epileptogenic potential in a zebrafish model system

As we were able to confirm the presence of 2-OPP in CSF and brain extracts of PDE-ALDH7A1 patients and *Aldh7a1* KO mice, we were interested to follow-up on the hypothesis that this metabolite may contribute to the epilepsy phenotype in patients. Towards this end, we subjected zebrafish larvae to different concentrations of 2-OPP and compared induced behavior to that after exposure to pentylenetetrazole (PTZ), a positive control compound known to induce epilepsy-like behavior in zebrafish (33).

Measurement of baseline behavioral activity of the zebrafish larvae 5 days post fertilization (dpf) revealed that 2-OPP significantly increased baseline activity at a concentration of 10 mM in the swimming water (Figure 7A). This effect of increased locomotor activity was notable at a concentration of 1 mM, however, statistical significance was only reached at 10 mM. All larvae survived for at least 24 hours at the tested doses, indicating that this effect does not result from immediate toxicity. To investigate if 2-OPP induces seizure-like hyperactivity behavior at 10 mM, zebrafish larvae were subjected to a previously published behavioral paradigm of light-dark cycles (34). After the addition of 2-OPP to the swimming water, larvae were allowed to acclimate in the behavioral chamber for 15 minutes, after which they were subjected to light-dark cycles. During the light-flashes, 72.5% of 2-OPP treated larvae displayed high-speed movements as compared to 37.5% of control treated larvae (Figure 7B). In addition, the cumulative duration of these high-speed movements was significantly increased upon treatment with 2-OPP. Further investigation of the observed hyperactivity behavior during the light-on periods and the entire period of light-dark cycles revealed that larvae treated with 10 mM 2-OPP had higher movement frequency and travelled longer distances during these movements, as compared to control treated larvae (Figure 7C-F). Typical seizure behavior, with whole-body convulsions culminating in loss-of-posture, was only observed in PTZ-treated larvae. However, the sign of hyperactivity upon 2-OPP treatment is in line with the seizure-like hyperactivity that is described in the genetic zebrafish PDE model (*aldh7a1* mutant) (34). This mutant displayed the first signs of seizure-like convulsions at 11 days post-fertilization, indicating that a certain build-up of metabolites might be required to induce this behavior.

**Figure 7.**
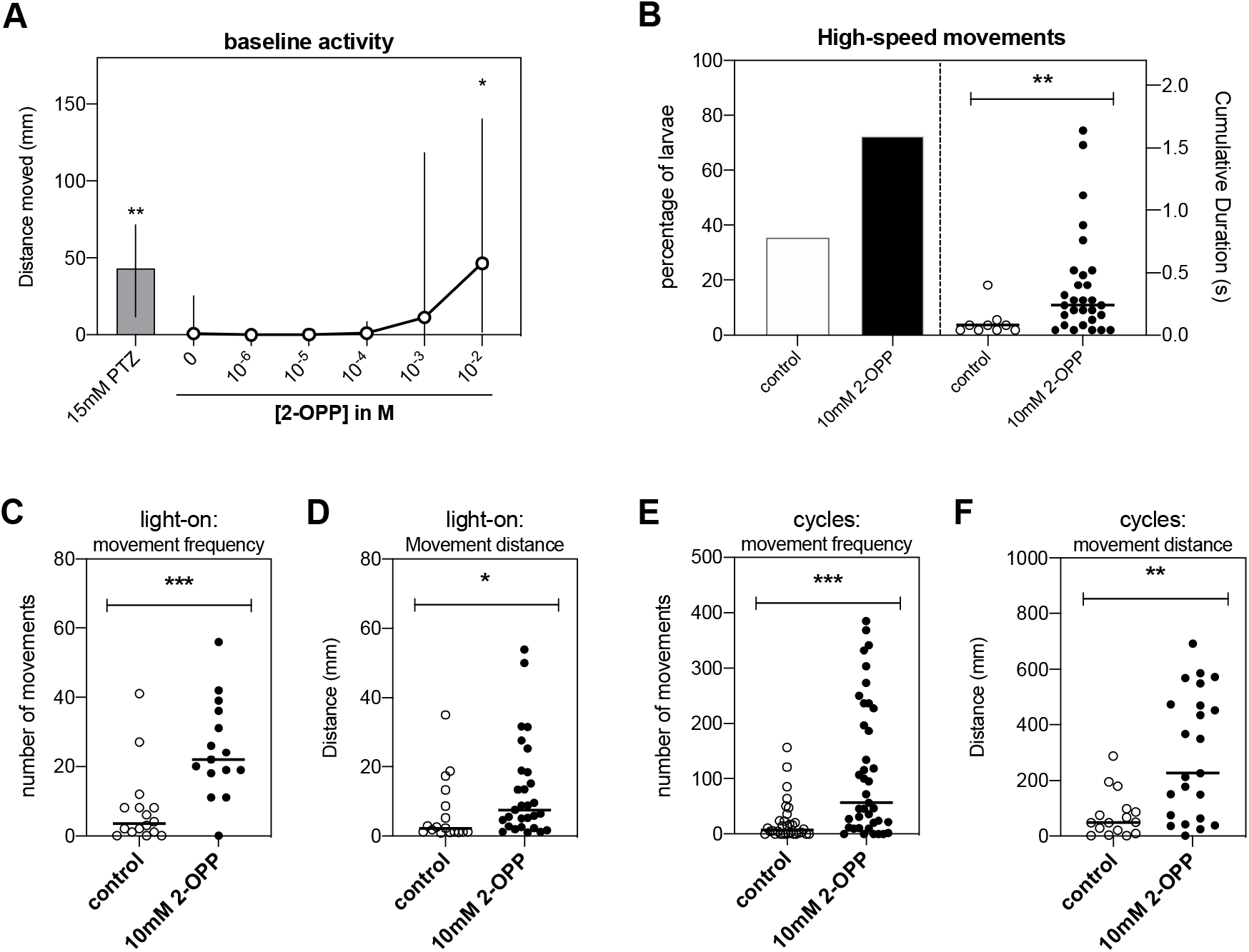
Hyperactivity behavior observed in zebrafish upon 2-OPP exposure. (**A**) Distance moved by 5 dpf zebrafish larvae after 10 min treatment with different concentrations of 2-OPP. PTZ was included as positive control of chemically-induced seizure-like hyperactivity. * = p<0.05, Kruskal Wallis test. (**B**) The percentage of zebrafish larvae that display high-speed movements after exposure to a series of light-flashes increased upon treatment with 10 mM 2-OPP (left y-axis). The duration of the high-speed movements was also significantly increased by 2-OPP (right y-axis). (**C-F**) The frequency of all movements, and the distance that the larvae moved was significantly increased during the light-on period (**C, D**) and during the entire 5-minute period of the light-flash paradigm (**E, F**), indicating that the observed increase in activity does not solely result from an increased response to the light-flash, but is indicative of hyperactivity throughout the light-flash period. For B-F: * = p<0.05, ** = p<0.01, *** = p<0.001, Mann-Whitney U test.

### Applicability of 2-OPP as a marker for PDE-ALDH7A1 in newborn screening

To set the first steps towards newborn screening for PDE-ALDH7A1, we also evaluated whether 2-OPP and 6-oxoPIP could be detected in DBS through a direct-infusion MS method without derivatization. This MS method was developed to be compatible with reagents commonly used for neonatal screening (Perkin Elmer, Neobase ™ 2 Non-derivatized MS/MS kit, which is for example used in the Dutch screening program). During the development of this direct-infusion MS method for DBS, it was apparent that for 6-oxoPIP, isobaric compounds were present in control DBS that prevented adequate distinction of PDE-ALDH7A1 patients from controls, while for 2-OPP, this was not the case (Supplemental Figure 5). Using the ratio between 2-OPP and hexanoyl/C6-carnitine, a marker already included in the Neobase kit, as a read-out, we could clearly distinguish between four PDE-ALDH7A1 patients tested and non-PDE controls (Figure 8). Of note, two of these PDE DBS were actual neonatal screening samples from PDE-ALDH7A1 patients, and had been stored for 12 to 13 years at room temperature before the current analysis, indicating high stability of 2-OPP as a DBS biomarker for PDE-ALDH7A1.

**Figure 8.**
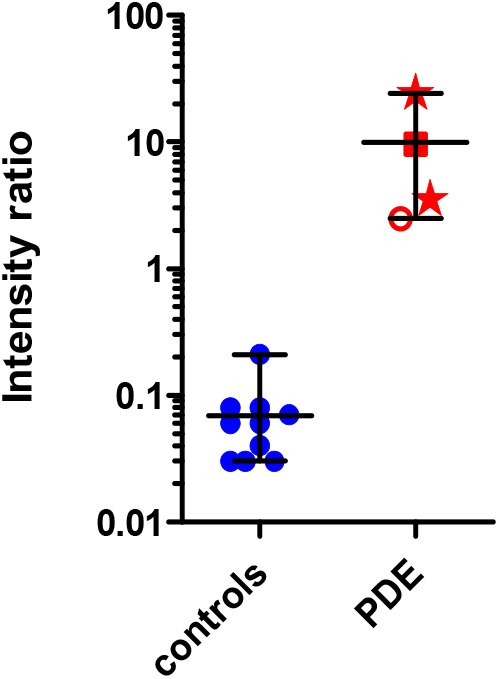
2-OPP based diagnosis of PDE-ALDH7A1 in DBS using a direct-infusion MS method. Y-axis shows relative intensity ratio between 2-OPP and C6-carnitine, while on the X-axis the categories of non-PDE controls (controls, N=10) and PDE-ALDH7A1 patients (PDE, N=4) are plotted. Patient results are marked as follows: Stars: neonatal DBS, untreated patients; filled square: DBS from child with PDE on vitamin B6 supplementation; open circle: DBS from untreated adolescent patient.

## Discussion

In this study, the power of combining untargeted metabolomics with infrared ion spectroscopy (IRIS) for the identification of novel diagnostic biomarkers is illustrated. We applied this innovative technology to address the clinical need for novel biomarkers for the treatable metabolic epilepsy syndrome PDE-ALDH7A1 to enable its inclusion in newborn screening. This has allowed for the identification of 2-OPP and 6-oxoPIP as novel PDE-ALDH7A1 biomarkers accumulating in plasma, urine, CSF, and brain tissue. We have developed a quantitative LC-MS/MS assay for diagnostic application of 2-OPP and 6-oxoPIP according to ISO:15189 criteria. Additionally, we have shown applicability of 2-OPP as a PDE-ALDH7A1 biomarker in DBS in a direct-infusion MS method compatible with newborn screening programs.

Regarding the biochemical relevance of 2-OPP in PDE-ALDH7A1 pathology, we have obtained initial evidence for epileptogenic potential of this compound in a zebrafish model system. Even though the tested concentration of 2-OPP in the swimming water that induced epilepsy-like behavior in zebrafish larvae was significantly higher than the concentration measured in body fluids and brain extracts of PDE-ALDH7A1 patients, it is likely that the actual 2-OPP concentration in the zebrafish brain was much lower than the concentration added to the swimming water. In line with this, 2-OPP intensity measured with NGMS in full-body lysates of 2-OPP exposed zebrafish larvae were 10^4^ fold lower as measured in the human brain sample (data not shown). As we have shown 2-OPP accumulation both in human patient and knock-out mouse brain tissue, it could be speculated that 2-OPP exerts neurotoxic effects on the brain and may in turn contribute to the ‘ongoing disease’ phenotype in treated PDE-ALDH7A1 patients. However, this hypothesis requires further experiments to determine the exact mechanism of action of 2-OPP on the brain, as well as how concurrent accumulation of α-AASA and P6C contributes to ongoing toxicity.

Of note, 2-OPP and 6-oxoPIP levels lacked a clear correlation with treatment status or patient genotype. In urine, overall a positive correlation between 2-OPP and 6-oxoPIP concentrations and levels of the known biomarker α-AASA was apparent. For most human plasma and urine samples obtained in an untreated state, the concentrations of 2-OPP and 6-oxoPIP were lower than those observed in patients treated solely with vitamin B6. This likely indicates that under treatment with vitamin B6 alone, substantial flux through the lysine pathway is ongoing and continues to lead to the formation of P6C-conjugates such as 2-OPP. Unfortunately, CSF samples from untreated patients were not available for analysis and comparison to the other body fluids, but it was apparent that also in CSF, highest 2-OPP and 6-oxoPIP levels were measured in patients on vitamin B6 monotherapy. Remarkably, 2-OPP levels in CSF were lowest in patients on B6 supplementation combined with arginine supplementation and in patients on triple therapy. Supplementation with arginine, which is thought to compete with lysine for transport across the blood-brain barrier, therefore seems to show a trend in lowering 2-OPP levels in CSF. Whether this biochemical effect in CSF correlates with clinical improvement upon arginine supplementation and lysine-reduction therapy in general, requires measurement of different patient samples in a larger, clinically well-defined PDE-ALDH7A1 patient cohort. In a subsequent study, we plan to evaluate the possible added value of 2-OPP (and 6-oxoPIP) as biomarkers for the clinical follow-up of PDE-ALDH7A1 patients. Also, we identified additional unknown signals in the untargeted metabolomics data which were significantly increased in PDE-ALDH7A1 patient body fluids. We will continue the process of identification of these unknown features to possibly yield additional insights on the biochemical consequences of α-AASA dehydrogenase deficiency. Of note, upon measurement of the 2-OPP model compound in the HPLC/electrochemical system in which previously an unknown compound (‘Peak X’) was detected in CSF of PDE patients by Hyland et al. (35, 36), it was apparent that this ‘Peak X’ could not be identified as 2-OPP (data not shown), indicating that the identity of ‘Peak X’ may still be waiting to be uncovered from our untargeted metabolomics data.

Another interesting finding from our study, with possible clinical implications, is the proposed mechanism of 2-OPP formation from the reaction of accumulating P6C with the ketone body acetoacetate. This would imply that especially in a state of ketosis, the increased concentration of acetoacetate would further drive 2-OPP formation in PDE-ALDH7A1 patients. As was stated in the introduction, in more than one third of patients, break-through epilepsy occurs during intercurrent (febrile) illness, for example gastroenteritis, despite increasing vitamin B6 dosage (15, 16). Catabolism often occurs during such episodes and, as is also known for other IEMs, this can lead to increased flux through amino acid metabolism as well as to ketosis. Even though we have not measured patient samples taken during such an intercurrent illness or period of reduced food intake, we speculate that increased formation of 2-OPP due to ketosis may trigger break-through seizures, despite of increased vitamin B6 dosage. Follow-up studies are required to further substantiate this hypothesis, and underpin the importance of an emergency plan for PDE-ALDH7A1 patients. Such an emergency plan prescribes increased caloric intake and protein reduction during intercurrent illness to avoid a catabolic situation. In the very recently updated consensus guidelines for the diagnosis and management of PDE-ALDH7A1 (37), it is recommended to double the vitamin B6 dosage for a maximum of three days and maintain caloric intake to prevent catabolism during times of illness. However, the level of evidence for this latter recommendation was scored low, as it was only based on expert opinion and could not be strengthened by underlying evidence. Our data may now provide the first biochemical evidence to further support this clinical recommendation. Also, these findings would again emphasize the importance of early diagnosis, as a ketogenic diet, which is a relatively common treatment in epilepsy, would be contra-indicated in PDE-ALDH7A1 patients.

Regarding the mechanism of formation of 2-OPP from P6C and acetoacetate, our incubation experiments point to a possible non-enzymatic process as this reaction also takes place in water and PBS. The only apparent difference with the reaction in plasma was the observed ratio between 2S,6S-2-OPP and 2S,6R-2-OPP; in the plasma incubation both enantiomers appeared, while in other matrices the 2S,6S enantiomer was formed preferentially. While the precise mechanism underlying the specific stereochemistry involved in 2-OPP formation will require further study, variability in the 2-OPP enantiomer ratio will not interfere with diagnostic interpretation of results in plasma, urine or CSF, as in these assays the sum of both 2-OPP enantiomers is used. Also for the DBS assay, the enantiomer ratio will not lead to diagnostic issues as both enantiomers are detected simultaneously in the direct-infusion MS method, which does not involve LC separation.

In conclusion, our research has uncovered 2-OPP and 6-oxoPIP as diagnostic PDE-ALDH7A1 biomarkers and has also provided the first evidence for practical applicability of 2-OPP as a biomarker in newborn screening. To our knowledge, our work represents the first example worldwide of the successful application of untargeted, high resolution MS metabolomics in combination with IRIS for the identification of a new biomarker for newborn screening of a treatable neurometabolic disorder. Although whole exome sequencing has been proposed as an alternative method for newborn screening, which may partly replace separate biochemical assays, a recent study emphasizes the crucial importance of biochemical testing to reach an acceptable level of sensitivity and specificity for newborn screening (38). To further confirm diagnostic specificity and sensitivity of 2-OPP as a PDE-ALDH7A1 biomarker, an extensive clinical validation study should be initiated as a next step, screening thousands of anonymized control neonatal DBS and additional samples from patients. Based on our current results, we are now ready to take on this next step to bring PDE-ALDH7A1 into newborn screening. Additionally, our results pose potential clinical implications regarding the importance of avoiding ketosis in PDE-ALDH7A1 patients. We will further explore these novel insights in future studies together with the international PDE Consortium and the PDE registry, ultimately to reach optimal clinical outcomes for all PDE-ALDH7A1 patients through early diagnosis and personalized biochemical and clinical follow-up.

## Methods

### Sample collection

Heparin-anticoagulated plasma, urine, CSF and/or DBS of human PDE-ALDH7A1 patients had been previously collected for routine metabolic screening or treatment follow-up. All patients (or their guardians) approved of the possible use of their leftover samples for method validation purposes, in agreement with institutional and national legislation, as reviewed by the accredited Research Ethics Committee of Radboud University Medical Centre (File number 2021-7296). For all PDE patients included, the diagnosis had been genetically confirmed previously. Control samples were obtained from leftover material from non-IEM patients who approved of the use of their anonymized leftover material for validation purposes, and which did not show abnormalities in targeted metabolite analyses. All plasma, urine and CSF samples were stored in a digital alarm controlled freezer at -20°C before analysis, for a period ranging from nine years to one month. The DBS were stored at room temperature before analysis; neonatal DBS were stored 12-13 years, the spot from an untreated adolescent patient was kept for 1.5 year at room temperature before analysis, while the spot of the vitamin B6-treated child was stored 1 month before analysis. Non-PDE control DBS had been stored at room temperature for a period between eight and twenty months.

With regard of the human brain tissue samples, full thickness samples of the cerebral cortex from a 9 year old female child with PDE-ALDH7A1 were taken at autopsy 12 hours postmortem and placed at −80° C. Clinical details on this patient were previously described (18, 31). Control brain tissue was obtained from the NICHD Brain and Tissue Bank for Developmental Disorders at the University of Maryland, Baltimore, MD, USA. Cortical tissue was also obtained from epilepsy surgery procedures. After excision, the fresh tissue was frozen in liquid nitrogen and stored at −80° C. Informed consent for the use of the tissue for research purposes was obtained from each subject or their legal guardian. For preparation of cortical extracts, frozen cortical tissue (100–200 mg) was separated from underlying white matter and homogenized in 5 mM Tris/HCl (pH 7.4) with 0.32 M sucrose. The homogenates were centrifuged at 3000 g for 5 min at 4°C, and the resulting supernatant was then centrifuged at 40000 g for 1 h at 4°C to pellet cellular membranes. The supernatant from this centrifugation was used for subsequent sample preparation for NGMS analysis.

The *Aldh7a1* KO mouse model used in this study was generated in accordance with guidelines of the Canadian Council on Animal Care and under an approved protocol from the University of British Columbia Animal Care Committee (Animal Protocols # A15-0200, A15-0180, A14-0031 and A18-0117), and was previously published (32). For collection of body fluids and tissues from *Aldh7a1* KO and WT mice for metabolite analysis, mice were anesthetized by Avertin (2,2,2-tribromoethanol) injection or isoflurane inhalation. The animals were then dissected, and blood was withdrawn directly from the heart by needle and syringe. Blood was collected in heparin tubes and plasma was separated by centrifuging the blood tubes at 3000 rpm for 10-15 min. Next, brain tissue samples were harvested. All samples were snap-frozen with isopentane in dry ice promptly after collection. Tissue and plasma samples were stored in a -80 ° C freezer until shipment.

Extractions were performed for two *Aldh7a1* KO and two WT mice brain tissues for NGMS analysis. For each tissue, a region containing mainly the cerebellum was dissected, weighing approximately 1030 mg (range 1003-1078 mg). In an Eppendorf tube, 400 ul of MeOH was added and the samples were centrifuged for 30 minutes at 18600 g at room temperature. The tissues were then pulverized and centrifuged again for 20 minutes at room temperature. The resulting supernatant was used for subsequent sample preparation for NGMS analysis.

### Sample preparation

Frozen plasma, urine, CSF or tissue extract samples were thawed at 4 °C and mixed by vortexing. A sample aliquot of 100 µL was transferred into a 1.5 mL polypropylene microcentrifuge tube. Then, 400 µL ice cold methanol/ethanol (50:50 vol/vol) containing five internal standards (caffeine-d3 0,88 µmol/L, hippuric-d5 acid 0.22 µmol/L, nicotinic-d4 acid 0.88 µmol/L, octanoyl-L-carnitine-d3 0.22 µmol/L, L-phenyl-d5-alanine 0,44 µmol/L (all from C/D/N Isotopes, Pointe-Claire, Canada)) was added to each aliquot. The samples were thoroughly mixed on a vortex mixer for 30 seconds. The samples were incubated at 4 °C for 20 minutes after which they were centrifuged at 18600 g for 15 minutes at 4 °C. An aliquot of 350 µL of the supernatant was transferred into a 1.5 mL polypropylene microcentrifuge tube. Samples were dried in a centrifugal vacuum evaporator (Eppendorf, Hamburg, Germany) at room temperature. The samples were reconstituted in 100 µL of water containing 0.1 % (vol/vol) formic acid, vortexed for 15 seconds and centrifuged at 18600 g for 15 minutes at room temperature. An aliquot of 90 µL was transferred into 250 µL polypropylene autosampler vials. These samples were either placed in an autosampler operating at 4 °C for direct analysis or stored at -80 °C. Stored samples were thawed at room temperature and centrifuged at 18600 g for 15 minutes at room temperature before analysis.

### Next Generation Metabolic Screening (NGMS)

NGMS was performed as previously described (20). In short, analyses were performed using an Agilent (Santa Clara, CA, USA) 1290 UHPLC system coupled to an Agilent 6545 QTOF mass spectrometer, equipped with a dual electrospray ionization (ESI) source. Each sample was run in duplicate in both positive and negative ionization modes. A 2.0 µL aliquot of extracted plasma sample was injected onto an Acquity HSS T3 (C18, 2.1 x 100 mm, 1.8 µm) column (Waters, Milford, MA, USA) operating at 40 °C. Chromatographic separations were performed by applying a binary mobile phase system. Exact details of buffer composition and MS settings were as previously reported (20). Each analytical batch was composed of control samples, PDE-ALDH7A1 patient samples, analytical quality control (QC) samples and a validation plasma pool to check for integrity of the automated data analysis pipeline. An analytical run consisted of a maximum of 150 samples. Control and patient samples were analyzed in duplicate. To correct for possible run-order influence on signal intensities, these duplicates were analyzed in anti-parallel run-order, meaning that duplicates of the first patient sample were analyzed on first and last position in the analytical run, while duplicates of the last patient sample were analyzed in the two middle positions of the run. Eight random control plasma samples were injected at the start of each analytical batch in order to condition the analytical platform. For further details on the analytical QC procedure and requirements, please refer to Coene et al (20).

### NGMS data processing and statistics

The output files of the NGMS runs were aligned using the open access software package XCMS (39) in single job modus. Full description of XCMS parameters is available on request. Following alignment and feature extraction, features were annotated against the Human Metabolome Database (HMDB (40)) for putative metabolite identification. For all features, two-sided t-tests were performed to identify significantly altered features between an individual patient and controls. These steps of alignment, annotation and statistical testing were integrated in an in-house bioinformatic pipeline which was validated for diagnostic use against ISO 15189 standards. Because of the large number of features identified in an individual patient sample (∼10000), the Bonferroni procedure was used to correct for multiple testing to prevent false positives, i.e. features incorrectly marked as significantly different in a patient. Two types of t-tests were applied to compare the intensity of each feature present in an individual patient sample to the intensities observed in the control samples as previously described (20). Only features that were marked as significantly different by both t-tests after Bonferroni correction (p-value <0.05) were retained for further analysis. Significantly different features were compared between PDE-ALDH7A1 patient samples to select common differential features.

### IRIS MS

Separations for the IRIS experiments were performed using a Bruker Elute SP HPLC system consisting of a binary pump, cooled autosampler and column oven using the exact same separation method as described above for NGMS. Fractions containing the metabolites of interest were obtained by collecting the eluent on 96-well plates using a Foxy R2 fraction collector. For these experiments, an injection volume of 20 µl was used.

IRIS experiments were performed in a quadrupole ion trap mass spectrometer (Bruker, AmaZon Speed ETD) modified for spectroscopy. Details of the hardware modifications and synchronization of the experiment with the infrared laser are described elsewhere (41). Collected LC-fractions and solutions of reference compounds (∼10^−7^ M in 50:50 methanol:water) were introduced at 80-180 μl/h flow rates to the electrospray source (+ESI). The ions of interest were mass-isolated and irradiated by the Free Electron Laser for Infrared eXperiments (FELIX) for IR analysis. IR spectra were recorded with FELIX set to produce IR radiation in the form of ∼10 μs macropulses of 50–150 mJ at a 10 Hz repetition rate (bandwidth ∼0.4% of the center frequency).

When the laser is resonant with a vibrational transition of the ions this leads to absorption of the IR photons, producing an increase in the ions’ internal energy and eventually leading to photodissociation. Thus, IR absorption can be observed by recording a fragmentation MS spectrum. IR spectra were constructed by plotting the fractional dissociation (IR yield ⍰= ⍰ΣI(fragment ions)/ΣI(parent + fragment ions)) as a function of IR laser frequency. The IR yield at each wavelength position of the laser was calculated from 4 to 8 averaged fragmentation mass spectra. The IR frequency was calibrated using a grating spectrometer, and the IR yield was linearly corrected for frequency-dependent variations in the laser pulse energy.

### P6C incubation experiments

P6C (from in house synthesis, see Supplemental Methods) was incubated in a 1 mM concentration (based on Mills *et al*. (3)) with 1 mM acetoacetate (See Supplemental figure 6 for synthesis of acetoacetate, not commercially available) for 24 hours at 37 °C in different matrices: H_2_O, PBS (pH 7.5), a non-IEM control plasma pool and a non-IEM control urine pool. These non-IEM control urine and plasma pools were prepared by mixing 4, respectively 7 anonymized left-over samples. Also, 1 mM P6C was incubated for 24 hours at 37 °C with a ketotic urine sample (based on increased excretion of 3-hydroxybutyric acid) and a ketotic plasma sample (based on increased concentration of acetyl-carnitine). After the 24 hour incubation period, samples were put on ice, and a 100 µL aliquot was taken for further processing for NGMS analysis, equal to the sample preparation procedure described above. The resulting samples were also used to record IR spectra according to the procedure described above.

### Zebrafish studies

All experiments were carried out in accordance with European guidelines on animal experiments (2010/63/EU). Tupfel long fin zebrafish were bred and raised in recirculation systems (temperature: ∼27° C, pH: 7.5–8, conductivity: ∼320 µS/cm) under a 14h:10h light-dark cycle. Fish were fed twice daily with Artemia sp. and Gemma Micro 300 (Skretting, Stavanger, Norway). Zebrafish eggs were obtained from natural spawning, and raised in E3 medium (5 mM NaCl, 0.17 mM KCl, 0.33 mM CaCl_2_, 0.33 mM MgSO_4_ and 1.3×10^−5^ % methylene blue) at 28.5° C until experiments.

At 5dpf the zebrafish were placed individually in a 96-well plate containing 100 µl E2 medium (5 mM NaCl, 0.17 mM KCl, 0.33 mM CaCl_2_ and 0.33 mM MgSO_4_). Directly before larvae were subjected to behavioral analyses, PTZ or 2-OPP were added to the wells via 100 µl of a 2x stock solution of compound in E2 medium. Videotracking of larval behavior was performed in a DanioVision system and tracked with EthoVision XT14 software from Noldus Information Technology (Wageningen, the Netherlands) at 28.5°C. The seizure-inducing protocol (34) consisted of 15 minutes of dark adaptation, followed by 5 cycles of 10 seconds light (100%) and 50 seconds darkness. After tracking, the 96-well plate was placed back in the incubator and subjected to the same behavioral paradigm after 24h of compound treatment. Behavioral responses were analyzed with EthoVision XT14 software, using start- and stop velocities of respectively 7.5 mm/sec and 5 mm/sec to determine movement frequencies, and a lower limit of >20 mm/sec to determine high-speed movements.

### Synthesis of ^13^C isotope labeled 6-oxoPIP and (2S,6R)-2-OPP

To enable quantitative mass spectrometry, ^13^C isotope labeled 6-oxoPIP and (2S,6R)-2-OPP derivatives were prepared, carrying one or two isotopic labels, respectively (Figure 9). Briefly, (*S*)-2-aminohexanedioic-6-acid carrying a single ^13^C label was dehydrated to afford an isotope labeled 6-oxoPIP analogue. The (2*S*,6*R*)-2-OPP isotope labeled standard was prepared by reacting a protected P6C derivative with ^13^C labeled acetone, followed by deprotection to afford the isotope labeled derivative of (2*S*,6*R*)-2-OPP (see Supplemental Figure 6 and 7 and the Supplemental Methods for more details on the synthesis).

**Figure 9.**
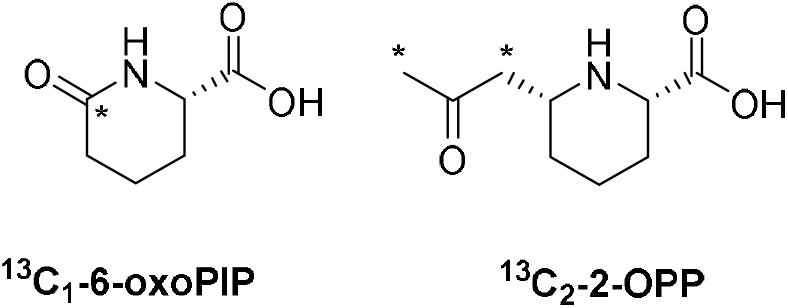
Isotope labeled standards of 6-oxoPIP and 2-OPP synthesized for this study; * denotes position of ^13^C.

### Quantitative LC-MS measurement of 6-oxoPIP and 2-OPP in body fluids

Levels of 6-oxo-PIP and 2-OPP were determined in plasma, urine and CSF using liquid chromatography tandem mass spectrometry (LC-MS) through a stable isotope dilution (SID) method. 10 µl of body fluid (for urine an equivalent to a kreatinine level of 0.1 mM) was diluted with 10 µl of stable isotope solution (5 µM of 1,3-^13^C_2_-(2S,6R)-2-OPP, respectively, 0.5 µM of ^13^C_1_-6-oxoPIP in H_2_O, see Figure 9 and Supplemental Information for more details), and 200 µl of H_2_O. For plasma, SID was carried out in a 30 KdA centrifugal filter (Amicon® Ultra 0,5 mL 30Kda centrifugal filter regenerated cellulose 30000 MWCO, Millipore) to remove protein (15 min. 14,000×g, 15°C). 1 µl of the isotope diluted sample was injected into the HPLC-MS/MS system, which consisted of a Waters I-Class Acquity fitted with a Atlantis T3 (2.1*100 mm dp 3µ) column connected to a Waters Xevo TQSµ mass spectrometer. The column was run at 400 µl/min in gradient mode using 0.5% acetic acid in H_2_O and acetonitrile (initial conditions 100% 0.5% acetic acid in H_2_O, gradient: 7.5% acetonitrile per minute) The column flow was directed to the Xevo TQSµ fitted with an electrospray ionization probe operating in the positive mode at unit resolution. The capillary voltage was set at 0.35 kV. The temperature settings for the source and ion block were respectively 550°C and 150°C. As drying gas, nitrogen was used at a flow rate of 800 L/h. The cone gas flow was set at 50 L/h. The collision cell was operated with argon as collision gas at a pressure of 0.35 Pa. For 6-oxoPIP, and its stable isotope, a neutral loss of the carboxylic group as -HCOOH was monitored (m/z 144.1 > m/z 98.1). For 2-OPP, and its stable isotope, the loss of the oxo-propyl group was monitored (m/z 186.1 > m/z 128.1). Quantification was done by comparison of the generated area response using the corresponding stable isotope as internal standard. This method was validated according to ISO:15189 standards for use in clinical diagnostic procedures.

### Direct infusion-MS measurement of 6-oxoPIP and 2-OPP in DBS

Analysis of 6-oxoPIP and 2-OPP in DBS was performed using a direct infusion tandem-MS method, similar to the Neobase ™ 2 non-derivatized MS/MS kit protocol. In short, DBS punches with a diameter of 6.35 mm were extracted with 300 µl MeOH containing internal standards. Extraction was performed by shaking at 45°C at 100 rpm for 30 minutes. After a 30 minutes cooling period, 50 µl of the extract was diluted with MeOH and 0,2% formic acid in H_2_O at a ratio of 1:1:0.2. Using a Waters I-Class Acquity HPLC-system, 10 µl of the diluted extract was infused into a Waters Xevo TQSµ at 40 µl/min using 80% acetonitrile and 20% 0.2% formic acid in H_2_O. The Xevo TQSµ which was set to monitor not only mrm transitions already part of the NeoBase protocol, but also the transitions 6-oxoPIP (m/z 144.1 > m/z 98.1) and 2-OPP (m/z 186.1 > m/z 128.1). For every compound, optimized cone and collision energies were used. The Xevo TQSµ was fitted with an electrospray probe and was operated at unit resolution. The capillary voltage was set at 3.0 kV. The temperature settings for the source and ion block were respectively 250°C and 150°C. As a drying gas, nitrogen was used at a flow rate of 250 L/h. The cone gas flow was set at 50 L/h. The collision cell was operated with argon as the collision gas at a pressure of 0.35 Pa. MS/MS response for 2-OPP and 6-oxoPIP was normalized on the response for hexanoyl (C6)-carnitine, a metabolite that is already analyzed in the Neobase ™ 2 kit. Hexanoyl-carnitine levels in control DBS were comparable to control levels for 2-OPP in DBS and had a narrow control range, making hexanoyl-carnitine optimally suited for ratio-based normalization.

## Data Availability

Raw data from metabolomics and IRIS experiments has been stored on institutional, backed-up servers and is available upon request.

## Author contributions

KLMC, JM, JO and RAW conceptualized the study. UFHE, KLMC, RAW, LAJK, MCDGH and TMAP performed data interpretation of untargeted metabolomics data. RvO, FAMGvG, GB, JM and JO were responsible for IRIS-MS experiments and interpretation. JMer, TJB, JMec and FPJTR designed and performed synthesis of model compounds and incubation experiments. AvR designed and performed the targeted LC-MS/MS and direct-infusion MS experiments. HHA-S and BRL provided *Aldh7a1* KO mice tissues and bodyfluids. Zebrafish studies were performed by SB, EdV and EvW. LAJ and SMGJr prepared human PDE and control brain tissue extracts. EAS performed targeted analysis of α-AASA in urine. KH performed analysis of model compounds with HPLC/electrochemical analysis. LAT analyzed data from the PDE registry. LAT, SM-A, MAAPW, LAB and CDMvK were involved in clinical management of PDE-ALDH7A1 patients and translation of results to clinical implications. PK provided bioinformatic support for untargeted metabolomics data analysis pipelines. UFHE and KLMC wrote the initial draft of the manuscript. All authors critically revised the manuscript and gave final consent for its submission.

## Acknowledgements

This research was partly funded by a ‘Stimuleringsbeurs’ from the Society for Inborn Errors of Metabolism for Netherlands and Belgium (ESN), a catalyst grant from United for Metabolic Disease (UMD-CG-2020-004), and a Stofwisselkracht grant under the project name ‘Innovative diagnostics in cerebrospinal fluid of patients with neurometabolic disorders’ (all to KLMC). Also, parts of this work were financially supported by an Interfacultary Collaboration Grant from Radboud University Nijmegen (to KLMC, RAW, JO and JM). The authors also gratefully acknowledge the Nederlandse Organisatie voor Wetenschappelijk Onderzoek (NWO), division Natural Sciences, for the support of the FELIX Laboratory (grant numbers VICI 724.011.002, TTW 15769, TKI-LIFT 731.017.419, Rekentijd 2019.062, all to JO). This work was also supported by an ERC-Stg grant (GlycoEdit, 758913) awarded to TJB. We are indebted to Siebolt de Boer, Joris Reintjes and Ed van der Heeft for technical assistance, and to Dawn Cordeiro for assistance in CSF sample distribution. Also we would like to thank our colleagues from the newborn screening laboratory at Elisabeth Tweesteden Ziekenhuis (ETZ) Tilburg, The Netherlands, for helpful discussions on newborn screening methodology. This research made use of metabolomics infrastructure that is part of the NWO-funded Netherlands X-omics initiative, project 184.034.019.

## Notes

### Competing Interest Statement

The authors have declared no competing interest.

### Author Declarations

The patient material used in this study had been previously collected for routine clinical laboratory diagnostics. All patients (or their guardians) approved of the possible anonimized use of their leftover samples for laboratory method validation purposes, in agreement with institutional and national legislation. The Research Ethics Committee of the Radboud University Medical Centre has approved of this study and has determined that the work was carried out in accordance with the applicable legislation concerning reviewal by an accredited research ethics committee (file number 2021-7296). The use of the autopsy material of the PDE-ALDH7A1 patient was previously approved by the Institutional Review Board of Seattle Childrens Hospital (file number FWA00002443).

### Summary of Updates

Author details and affiliations were updated.

