## Supplemental Information for "Identification of novel biomarkers for pyridoxine-dependent epilepsy using untargeted metabolomics and infrared ion spectroscopy - biochemical insights and clinical implications"

^#^Corresponding authors

**Supplemental Figure 1.** Correlation of quantitative concentrations of 6-oxoPIP and 2-OPP in plasma (X-axis) with normalized NGMS intensity values (Y-axis, relative units). R^2^ from linear regression is shown.

| 6-oxoPIP (μmol/L) | Normalized Intensity 6-oxoPIP NGMS |
| --- | --- |
| 3.9 | 20.4164 |
| 9.4 | 35.1139 |
| 4.99 | 19.8523 |
| 11.45 | 46.5793 |
| 2.44 | 11.8292 |
| 7.62 | 54.283 |
| 6.21 | 55.9013 |
| 15.35 | 118.664 |
| 10.86 | 72.3872 |

| 2-OPP (R+S) (μmol/L) | Normalized Intensity 2-OPP (R+S) NGMS |
| --- | --- |
| 1.75 | 202.308 |
| 2 | 154.058 |
| 2.09 | 166.968 |
| 2.61 | 201.13 |
| 1.48 | 125.495 |
| 5.23 | 473.753 |
| 3.53 | 275.151 |
| 4.72 | 319.248 |
| 1.83 | 105.477 |

**Supplemental Figure 2.** Within-sample correlation for 2-OPP and 6-oxoPIP levels in urine, plasma and CSF of PDE-ALDH7A1 patients. While for urine, there appeared to be a positive correlation between 2-OPP and 6-oxoPIP levels, in plasma and CSF this was not clear. Different treatment regimens in patients are coded as follows: open circles: untreated; filled squares: vitamin B6 supplementation; filled circles: vitamin B6 and arginine supplementation; filled triangles: vitamin B6 supplementation and lysine restriction; filled diamonds: vitamin B6 and arginine supplementation and lysine restriction; cross: therapy unknown.

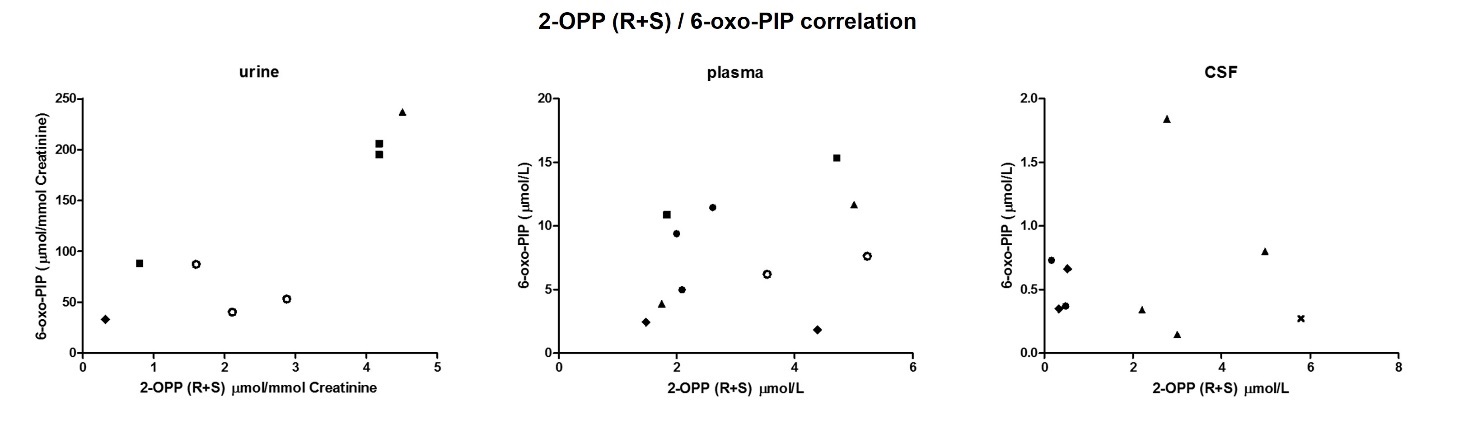

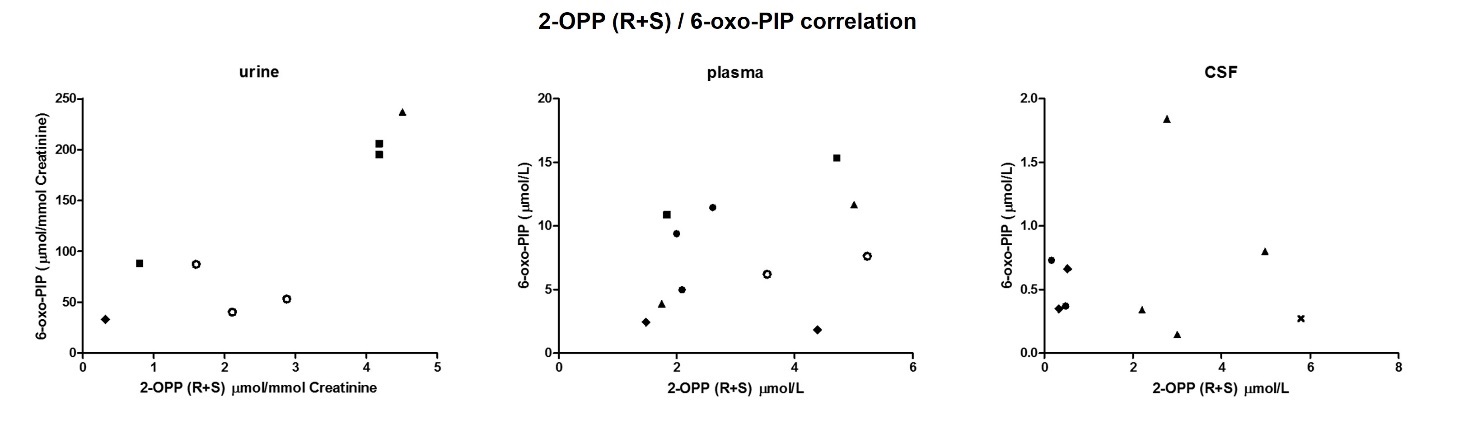

**Supplemental Figure 3.** Correlation of 6-oxoPIP and 2-OPP concentration to α-AASA concentration in urine of PDE-ALDH7A1 patients. R^2^ from linear regression is shown.

| α-AASA (μmol/mmol creatinine) | 6-oxoPIP (μmol/mmol creatinine) |
| --- | --- |
| 1.9 | 60.7 |
| 1.2 | 28.8 |
| 2.3 | 35.4 |
| 7.5 | 155 |
| 1.2 | 62.5 |
| 10.6 | 129 |
| 1.2 | 33.18 |
| 12.7 | 205.85 |
| 6.5 | 83 |

| α-AASA (μmol/mmol creatinine) | 2-OPP (R+S) (μmol/mmol creatinine) |
| --- | --- |
| 1.9 | 1.31 |
| 1.2 | 1.783 |
| 2.3 | 2.242 |
| 7.5 | 3.57 |
| 1.2 | 0.693 |
| 10.6 | 3.486 |
| 1.2 | 0.32 |
| 12.7 | 4.18 |
| 6.5 | 1.8 |

**Supplemental Figure 4.** (**A**) Overlay of IR spectra of the protonated 2S,6S-2OPP ion measured from incubations of P6C and AcAc in plasma, urine, H_2_O and PBS, incubation of P6C in ketotic urine and from a solution of the synthetic reference standard. (**B**) Overlay of IR spectra of the protonated 2S,6R-2OPP ion measured from incubations of P6C and AcAc in plasma and H_2_O and from a solution of the synthetic reference standard.

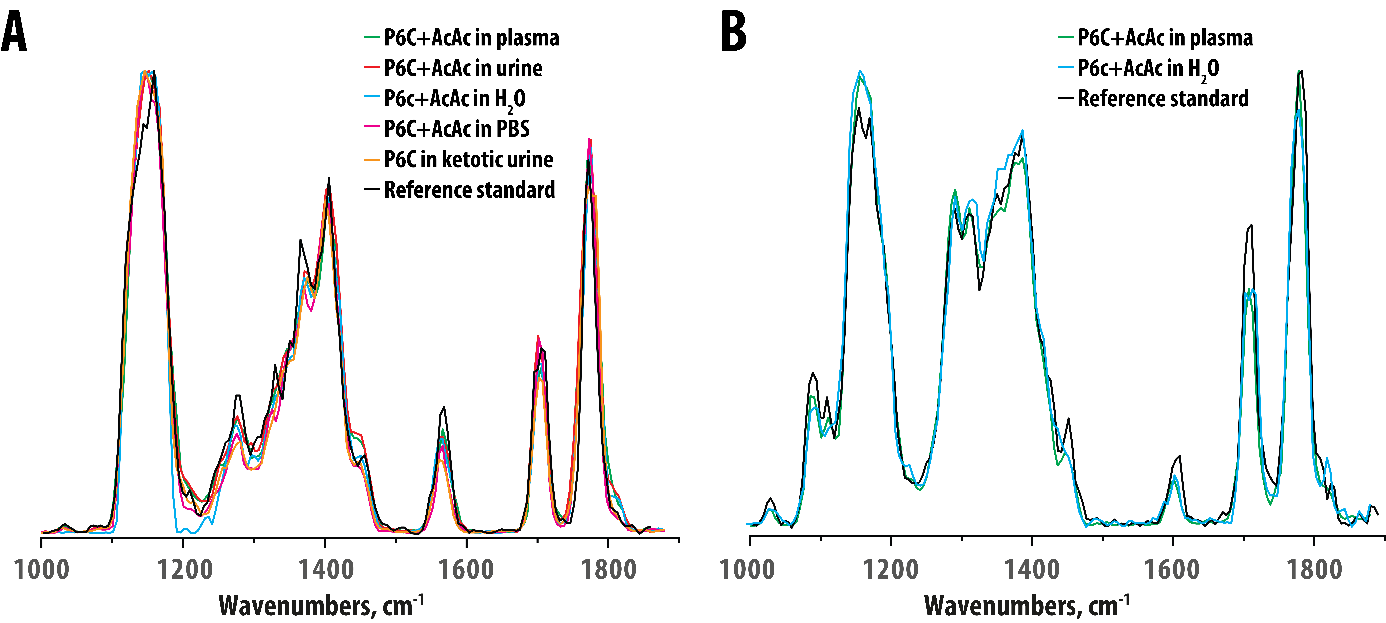

**Supplemental Figure 5.** Direct infusion MS results in DBS for 6-oxoPIP and 2-OPP, showing complete overlap between PDE patients and controls for 6-oxoPIP, but adequate distinction based on 2-OPP. Y-axis shows intensity of 6-oxoPIP in the left figure and 2-OPP in the right figure. On the X-axis, the categories of non-PDE controls (controls, N=10) and PDE-ALDH7A1 patients (PDE, N=4) are plotted. Patient results are marked as follows: Stars: neonatal DBS, untreated patients; filled square: DBS from 10-year old patient on vitamin B6 supplementation; open circle: DBS from untreated 16-year old patient.

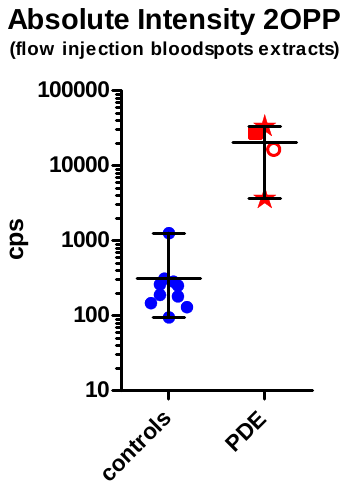

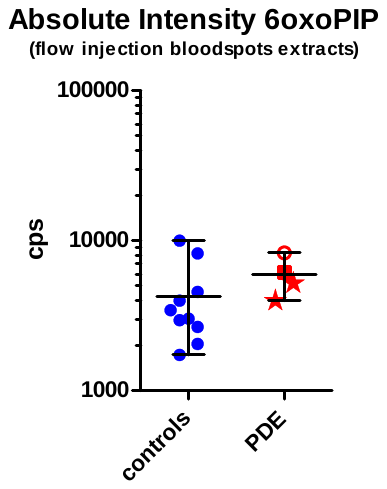

**Supplemental Figure 6.** Synthesis of 2-OPP, 1,3-^13^C_2_-(2*S*,6*R*)-2-OPP and ^13^C_1_-6-oxoPIP; reagents (**1-14**) and reaction conditions (**a-m**): (**1**) α-amino adipic acid, (**2**) (*S*)-6-oxopiperidine-2-carboxylic-6-^13^*C* acid (6-oxoPIP), (**3**) tert-butyl acetoacetate, (**4**) acetoacetate, (**5**) allysine ethylene acetal, (**6**) (*S*)-2,3,4,5-tetrahydropyridine-2-carboxylic acid (P6C), (**7**) benzyl (2*S*)-1-benzyl-6-(2-oxo-propyl) piperidine-2-carboxylate, (**8**) (2*S*)-6-(2-oxopropyl)piperidine-2-carboxylic acid (2-OPP), (**9**) acetone-1,3-^13^C_2_, (**10**) *tert*-butyldimethyl(prop-1-en-2-yloxy)silane ^13^C_2_, (**11**) benzyl 2-(((benzyloxy)carbonyl)amino)-5-(1,3-dioxolan-2-yl)pentanoate, (**12**) (*S*)-1-((benzyloxy)carbonyl)-1,2,3,4-tetrahydropyridine-2-carboxylic acid, (**13**) dibenzyl (2*S*)-6-(2-oxopropyl-1,3-^13^C_2_)piperidine-1,2-dicarboxylate, (**14**) (2*S*)-6-(2-oxopropyl-1,3-^13^C_2_)piperidine-2-carboxylic acid, (**a**) AcOH, 120 ^o^C, 95%; (**b**) TFA, DCM, 96%; (**c**) 1M HCl, quant; (**d**) H_2_O (pH 6-7); (**e**) BnBr, K_2_CO_3_, DMF, 40% over two steps; (**f**) H_2_, Pd/C, ACN, 54%; (**g**) TBDMSOTf, 0 ^o^C, DCM, 92%; (**h**) CbzOsu, NaHCO_3_, dioxane/H_2_O; (**i**) BnBr, K_2_CO_3,_ DMF, 85% over two steps; (**j**) *p-*TsOH, DMF, toluene, 115 ^o^C, 94%; (**k**) AcCl, MeOH, 99%; (**l**) Sn(OTf)_2_, MeCN, -30 ^o^C to room temperature, 79%; (**m**) H_2_, Pd/C, MeOH/H_2_O, 76%. The * denotes the position of the ^13^C isotope label. Please refer to supplemental methods for detailed description of synthesis.

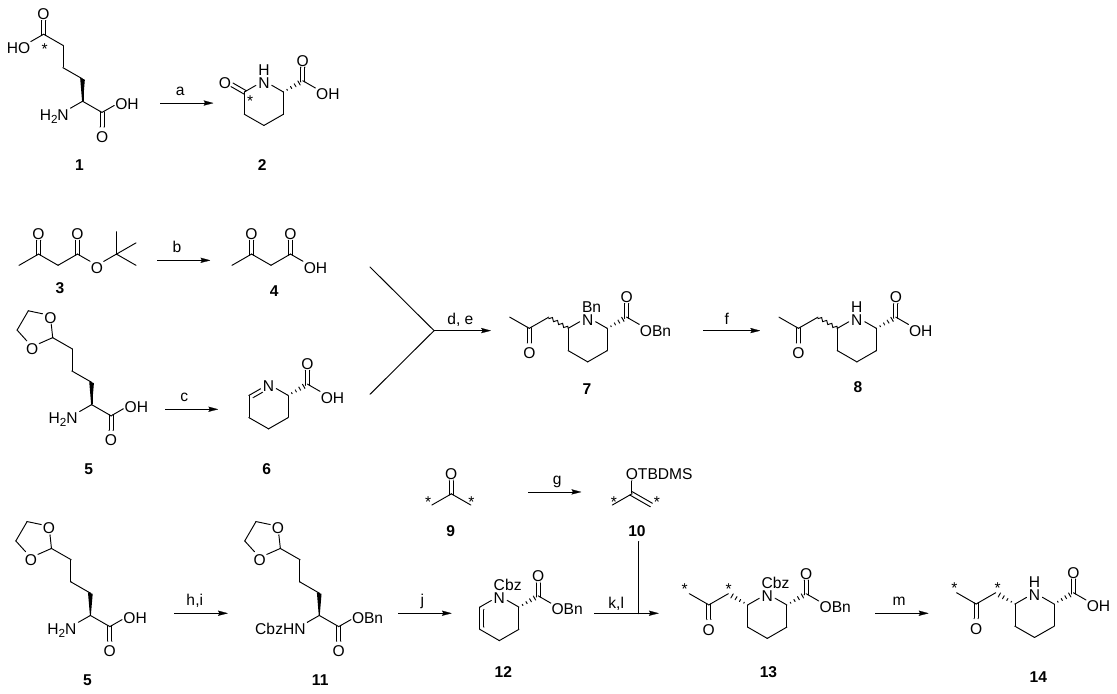

**Supplemental Figure 7.** ^1^H and ^13^C NMR spectra of compounds 2, 7, 8 and 10-14 as shown in Supplemental Figure 6. For further details on NMR spectra, please refer to Supplemental Methods, which are described below.

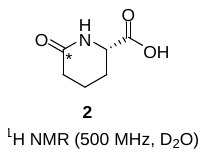

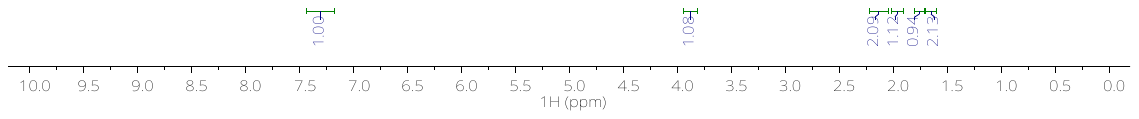

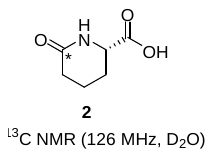

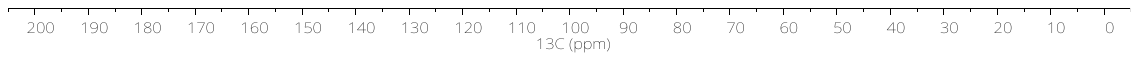

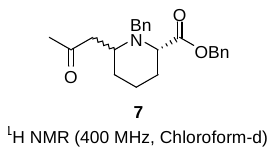

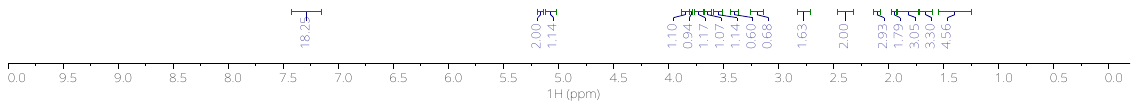

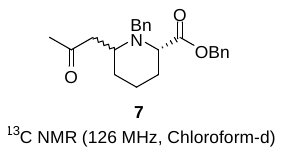

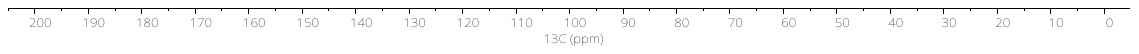

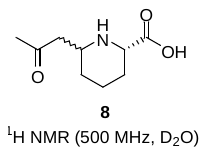

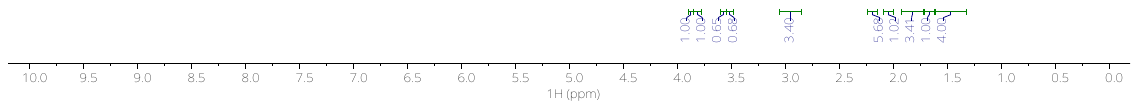

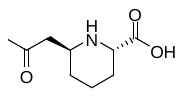

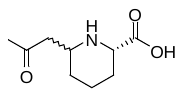

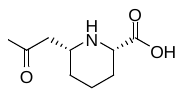

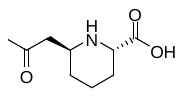

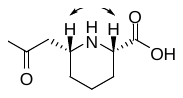

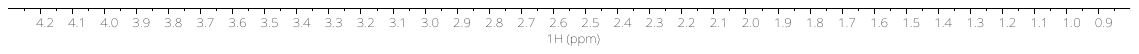

NOE (3.90 ppm)

^1^H (500 MHz, D_2_O)

TOCSY (3.60 ppm)

TOCSY (3.90 ppm)

NOE (3.60 ppm)

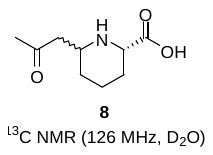

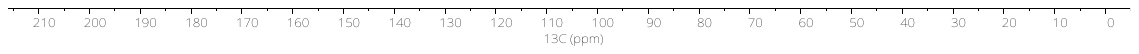

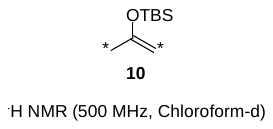

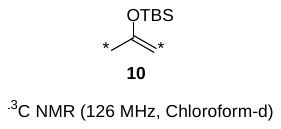

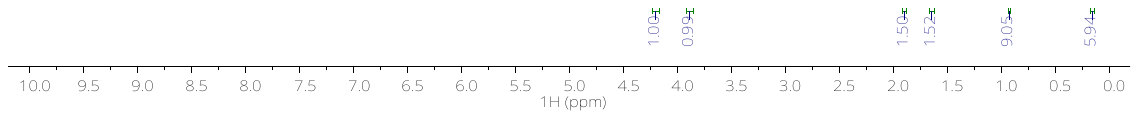

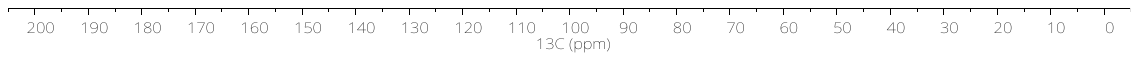

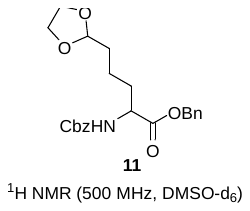

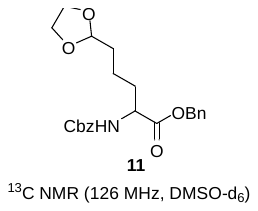

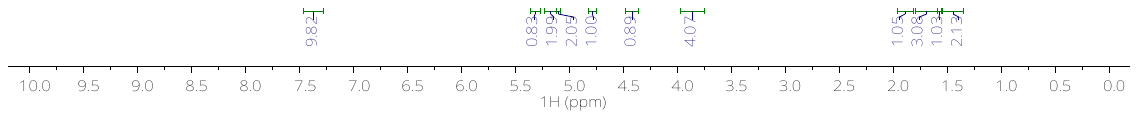

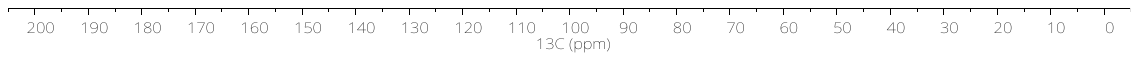

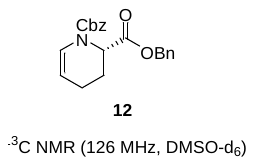

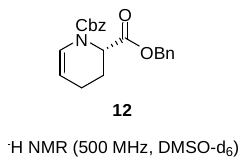

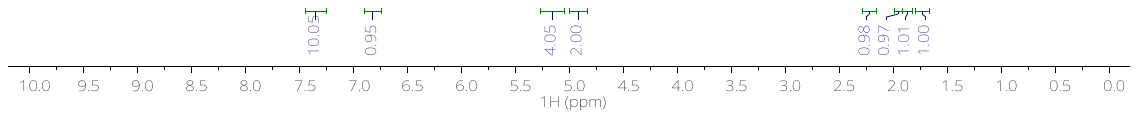

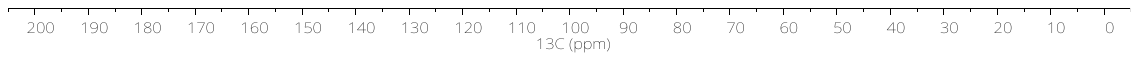

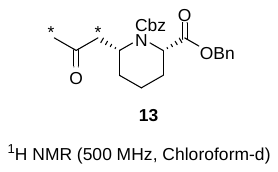

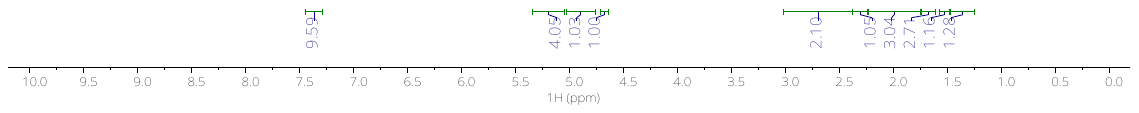

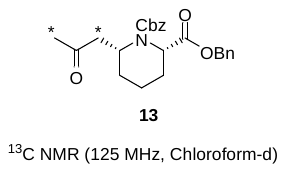

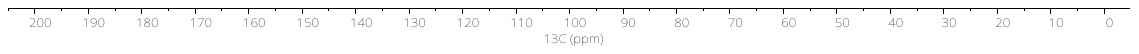

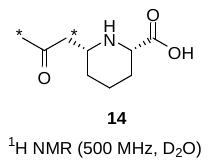

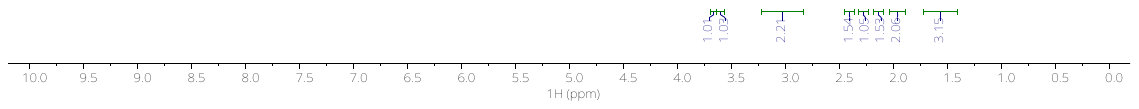

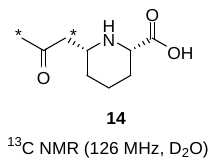

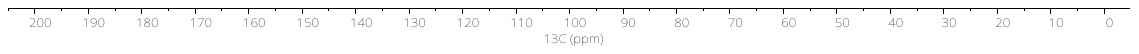

**Supplemental Methods**

^1^H and ^13^C NMR spectra and (LC) MS measurements in model compound synthesis

^1^H and ^13^C NMR spectra (Supplemental Figure 7) were recorded on a Bruker AVANCE III (500 MHz ^1^H, 125 MHz ^13^C) or) equipped with a Bruker Prodigy BB cryoprobe or a Bruker AVANCE III (400 MHz ^1^H, 100 MHz ^13^C in the solvent indicated at room temperature (RT). Chemical shifts are reported in *δ* (ppm) units relative to the internal reference tetramethylsilane (Me_4_Si). For ^1^H NMR spectra, the following abbreviations are used to describe multiplicities: s (singlet), d (doublet), t (triplet), bs (broad singlet), dd (double doublet), and m (multiplet). Coupling constants are reported in Hertz (Hz) as a *J* value, for ^13^C labelled compounds a distinction is made between ^1^J and ^3^J couplings. Mass spectra were recorded on Thermo Finnigan LCQ Advantage Max. LC-MS was carried out on a Shimadzu LCMS-QP8000 (Duisburg, Germany) single quadrupole bench-top mass spectrometer operating in a positive ionization mode. The scanning range was m/z 50-2000. A gradient of MeCN/H_2_O containing 0.1 % formic acid was used. Samples were injected using a flow rate of 0.2 mL/min and eluted with 5-100 % in 50 min, infused in the Electrospray system. All compounds were routinely checked by TLC on Kieselgel 60 F254 (Merck, Darmstadt, Germany); spots were visualized under UV light (254 nm) and were stained with ninhydrin, 2-4-ninitrophenylhydrazine (DNP), Cerium Molybdate Stain or aqueous KMnO_4_ (depending on the reaction), followed by heating on a hot plate. Rf values were obtained with the indicated solvent mixtures. All solvents were reagent grade and, when necessary, were purified and dried by standard methods. Organic solutions were dried over anhydrous sodium sulfate and anhydrous magnesium sulfate. The yields of the samples were calculated after drying the samples under high vacuum overnight or after lyophilization. All commercially purchased reagents were used without further purification as delivered from the corresponding companies. (*S*)-2-Aminohexanedioic-6-^13^*C* acid, *tert*-butylacetoacetate, Sn(OTf)_2_ and acetone-1,3-^13^C_2_ were obtained from Sigma Aldrich. Allysine ethylene acetal was obtained from Chiralix. TBDMSOTf and CbzOSU were obtained from Fluorochem.

Detailed information on synthesis as shown in Supplemental Figure 6, and characteristics for NMR spectra as shown in Supplemental Figure 7.

Numbers in brackets refer to compounds as shown in Supplemental Figure 6. Superscript numbers in parentheses refer to Supplemental References.

**(*S*)-6-Oxopiperidine-2-carboxylic-6-^13^*C* acid [2]** ^(1)^

(*S*)-2-Aminohexanedioic-6-^13^*C* acid (2.15 mg, 13.3 µmol) was dissolved in AcOH (200 µL) and refluxed for 6 h. The solution was cooled to RT, concentrated under reduced pressure, dissolved in water and lyophilized to obtain 1.81 mg of **2** (12.6 µmol, 95%) as a white solid. ^1^H NMR (500 MHz, DMSO-d6) δ 7.34 (s, 1H), 3.88 (qd, *J* = 5.6, 2.3 Hz, 1H), 2.19 – 2.06 (m, 2H), 2.03 – 1.89 (m, 1H), 1.78 – 1.70 (m, 1H), 1.69 – 1.59 (m, 2H); ^13^C NMR (126 MHz, DMSO-d6) δ 173.7, 55.0, 31.0, 25.2, 18.7.

**Acetoacetic acid [4]** ^(2)^

TFA (15 mL, 190 mmol) was added to an ice-cold solution of *tert*-butylacetoacetate (10 mL, 61 mmol) in DCM (15 mL). The solution was allowed to warm up after 5 min and continuously stirred for 1 h. The solution was concentrated *in vacuo* to obtain 6.0 g of acetoacetic acid in 96% yield (59 mmol), used as is without further purification. *R*_F_ = 0.30 (1:1, EtOAc:heptane).

**(*S*)-2,3,4,5-Tetrahydropyridine-2-carboxylic acid, P6C [6]**

For the incubations: To a suspension of allysine ethylene acetal (200 mg) in water (5 mL) was added 800 mg Amberlyst-15. After complete conversion the Amberlyst resin was washed with water (2 x 5 mL), subsequently the P6C was eluted with 10 mL 25% ammonia solution and dried *in vacuo* at ambient temperature.^(3)^ The resulting solid was reconstituted to a 1 mM solution used in the incubations.
For synthesis: allysine ethylene acetal (4.0 g) was dissolved in 1 M HCl (42 mL). After complete conversion the resulting solution was used as is for the synthesis of **7.**

**Benzyl (2*S*)-1-benzyl-6-(2-oxopropyl)piperidine-2-carboxylate [7]**

A solution of acetoacetic acid (**4**) (6.0 g, 59 mmol) in water (20 mL) was neutralized to pH 7 with solid NaOH. The acidic solution of **6** was added dropwise to the mixture while keeping the pH between 6 and 7 with addition of 1 M NaOH. After the solution (pH = 6.5) was stirred overnight it was refluxed for 30 min and subsequently concentrated *in vacuo*. Hot filtration with ethanol resulted in 3.9 g of a crude mixture containing **8.**^(4)^ To facilitate purification, the residue was derivatized. The resulting residue was redissolved in DMF (60 mL) and cooled to 0 ^o^C. Solid K_2_CO_3_ (15 g, 11 mmol) was added followed by the addition of BnBr (10 mL, 84 mmol). The resulting mixture was stirred overnight and concentrated *in vacuo*. The residue was taken up in EtOAc (50 mL), washed with water (10 mL) and brine (10 mL), dried over MgSO_4_ and concentrated *in vacuo*. Purification by column chromatography (3% diethyl ether in toluene) yielded 3.2 g of a slightly red oil in 42% yield (21 mmol) of both stereoisomers in a ratio of 1:0.6 (cis:trans, assignment of diastereomers based on NMR **8**). *R*_F_ = 0.29 (1:3, EtOAc:heptane); ^1^H NMR (400 MHz, CDCl_3_, both stereoisomers; integrals based on major stereoisomer) δ 7.50 – 7.03 (m, 18H), 5.13 (s, 2H), 5.05 (d, *J* = 2.1 Hz, 1H), 3.82 (d, *J* = 14.3 Hz, 1H), 3.77 (s, 1H), 3.75 – 3.67 (m, 1H), 3.62 (d, *J* = 14.3 Hz, 1H), 3.53 (t, *J* = 4.8 Hz, 1H), 3.38 (dd, *J* = 7.6, 4.3 Hz, 0.6H), 3.17 (s, 0.6H), 2.79 – 2.68 (m, 1.6H), 2.42 – 2.31 (m, 2H), 2.08 (s, 3H), 1.92 (s, 1.8 H), 1.90 – 1.71 (m, 3H), 1.70 – 1.57 (m, 3H), 1.52 – 1.24 (m, 4.5H); ^13^C NMR (126 MHz, Chloroform-d) δ 174.6, 173.2, 139.6, 128.5, 128.4, 128.30, 128.29, 128.23, 128.21, 128.17, 66.3, 66.1, 63.2, 60.1, 57.5, 55.6, 54.1, 52.2, 47.6, 46.7, 30.4, 30.2, 29.9, 28.4, 26.0, 20.32, 20.26; HRMS (ESI+); calcd for C_23_H_28_NO_3_ (*M*+H^+^): 366.2069, found 366.2081.

**(2*S*)-6-(2-Oxopropyl)piperidine-2-carboxylic acid [8, 2-OPP]**

A solution of **7** (623 mg, 1.70 mmol) in MeCN (10 mL) was purged with argon. Pd/C (181 mg, 0.17 mmol) was added and the atmosphere was exchanged to H_2_. After almost complete conversion (~ 1.5 h) the mixture was filtered over celite and concentrated *in vacuo*. The residue was purified by chromatography on Iatro beads (0 -> 40% MeOH in DCM) to yield 172 mg of an off white solid (55%, 0.93 mmol). ^1^H NMR (500 MHz, D_2_O, both diastereoisomers; integrals based on major stereoisomer) δ 3.96 (t, *J* = 5.0 Hz, 1H), 3.94 – 3.85 (m, 1H), 3.66 (dd, *J* = 12.5, 3.3 Hz, 0.65H), 3.61 (dtd, *J* = 12.3, 6.3, 2.9 Hz, 0.65H), 3.13 – 2.92 (m, 3.4H), 2.27 (s, 5.7H), 2.18 – 2.09 (m, 1H), 2.03 – 1.82 (m, 3.4H), 1.80 – 1.71 (m, 1H), 1.70 – 1.39 (m, 4H); ^13^C NMR (126 MHz, D_2_O, both stereoisomers) δ 211.9, 210.7, 174.3, 173.4, 60.2, 56.1, 52.3, 49.5, 45.5, 44.3, 29.6, 29.5, 27.6, 27.1, 26.2, 24.7, 22.1, 18.5; HRMS (ESI+); calcd for C_9_H_16_NO_3_ (*M*+H^+^): 186.1130, found 186.1130.

***tert*-Butyldimethyl(prop-1-en-2-yloxy)silane ^13^C_2_ [10]** ^(5)^

A flame dried flask was charged with dry DCM (10 ml) and acetone-1,3-^13^C_2_ (200 mg, 3.33 mmol). The solution was cooled with an ice-bath and TEA (550 µL, 4.00 mmol) was added. After 1 h at RT, the solution was cooled with an ice-bath and TBDMSOTf (841 µL, 3.66 mmol) was added dropwise. After stirring at RT for 4 h, the reaction was quenched with cold sat. aq. NH_4_Cl (5 mL). The mixture was extracted with Et_2_O (15 mL), dried over MgSO_4_ and concentrated under reduced pressure. The resulting non –viscous clear liquid was separated by means of pipet from the solids and viscous red oil resulting in 530 mg of **10** (3.06 mmol) in 92% yield. ^1^H NMR (500 MHz, chloroform-d) δ 4.24 – 4.20 (m, 1H), 3.93 – 3.90 (m, 1H), 1.77 (ddd, ^1^*J_CH_* = 126,7 Hz, *^3^J =* 3.9, 0.9 Hz, 3H), 0.95 (s, 9H), 0.18 (s, 6H); ^13^C NMR (126 MHz, Chloroform-d) δ 91.3, 91.2, 25.7, 22.8, 22.7, -4.6.

**Benzyl 2-(((benzyloxy)carbonyl)amino)-5-(1,3-dioxolan-2-yl)pentanoate [11]** ^(6)^

A suspension of (*S*)-2-amino-5-(1,3-dioxolan-2-yl)pentanoic acid (2.00 g, 10.6 mmol) in dioxane/water (10 mL, 1:1) was cooled with an ice bath and subsequently NaOH (444 mg, 11.1 mmol), NaHCO_3_ (1.33 g, 15.9 mmol) and CbzOSu (2.69 g, 10.8 mmol) were added. After stirring overnight, dioxane was removed under reduced pressure. The resulting solution was diluted with a 2.5% NaHCO_3_ solution (100 mL) and washed with Et_2_O (3 x 25 mL). The basic aqueous layer was acidified with 6M HCl solution until precipitation was observed (pH ~ 3). The resulting precipitate was extracted with EtOAc (3 x 50 mL), dried over MgSO_4_ and concentrated under reduced pressure. The resulting transparent oil was dissolved in dry DMF (20 mL) and cooled with an ice bath. Subsequently K_2_CO_3_ (1.53 g, 11.1 mmol) and BnBr (1.88 mL, 15.9 mmol) were added. The reaction mixture was stirred overnight at RT. After complete conversion water (100 mL) was added followed by extraction with EtOAc (3 x 50 mL). The organic layer was washed with brine (50 mL), dried over MgSO_4_ and concentrated under reduced pressure. The resulting residue was purified by column chromatography (heptane:EtOAc = 2:1) to yield 4.03 g of a transparent oil (10.6 mmol, 85%). *R*_F_ = 0.29 (1:3, EtOAc:heptane); ^1^H NMR (400 MHz, Chloroform-d) δ 7.39 – 7.28 (m, 10H), 5.29 (d, *J* = 8.2 Hz, 1H), 5.24 – 5.13 (m, 2H), 5.10 (s, 2H), 4.79 (t, *J* = 4.6 Hz, 1H), 4.43 (q, *J* = 7.7, 7.1 Hz, 1H), 3.98 – 3.75 (m, 4H), 1.97 – 1.83 (m, 1H), 1.79 – 1.60 (m, 3H), 1.47 (m, 2H); ^13^C NMR (126 MHz, Chloroform-d) δ 128.8, 128.7, 128.6, 128.4, 128.3, 128.2, 104.2, 67.3, 67.1, 65.01, 65.00, 54.1, 33.4, 32.6, 19.8; HRMS (ESI+): calcd for C_32_H_27_NNaO_6_ (*M*+Na^+^): 374.1368, found 374.1369

**(*S*)-1-((Benzyloxy)carbonyl)-1,2,3,4-tetrahydropyridine-2-carboxylic acid [12]** ^(6)^

To a solution of **11** (2.60 g, 6.3 mmol) in toluene (25 mL) was added DMF (240 µL, 3.14 mmol) and *p*TsOH·H_2_O (120 mg 0.63 mmol). The solution was heated to reflux for 3 h. Upon complete conversion the reaction was cooled to RT, diluted with EtOAc (25 mL) and quenched with saturated aqueous NH_4_Cl (25 mL). The layers were separated and the organic layer was washed with sat. aq. NaHCO_3_, dried over MgSO_4_ and concentrated under reduced pressure. The resulting residue was purified by column chromatography (EtOAc:heptane = 1:4) resulting in 2.10 g of a colorless oil in 94% yield (6.29 mmol). *R*_F_ = 0.45 (1:3, EtOAc:heptane); ^1^H NMR (500 MHz, DMSO-d6, rotamers) δ 7.40 – 7.24 (m, 10H), 6.84 – 6.76 (m, 1H), 5.23 – 5.06 (m, 4H), 5.01 – 4.81 (m, 2H), 2.34 – 2.16 (m, 1H), 1.95 (dt, *J* = 17.6, 5.7 Hz, 1H), 1.87 (dddd, *J* = 18.8, 10.1, 5.1, 2.1 Hz, 1H), 1.74 (m, 1H); ^13^C NMR (126 MHz, chloroform-d, rotamers) δ 170.4, 170.1, 152.6, 152.5, 163.13, 136.07, 135.8, 135.7, 128.5, 128.42, 128.36, 128.10, 128.09, 128.07, 127.9, 127.71, 127.70, 127.4, 124.1, 123.8, 105.8, 105.4, 67.1, 67.0, 66.34, 66.29, 55.7, 55.4, 23.13, 12.08, 17.9, 17.7; HRMS (ESI+): calcd for C_21_H_21_NNaO_4_ (*M*+Na^+^): 374.1368, found 374.1369.

**Dibenzyl (2*S*)-6-(2-oxopropyl-1,3-^13^C_2_)piperidine-1,2-dicarboxylate [13]**

To a solution of **12** (478 mg, 1.36 mmol) in dry MeOH (15 mL) was added AcCl (20 µL, 0.27 mmol) at RT. After stirring for 1h, the solution was diluted with DCM (5 ml) and quenched with saturated aqueous NaHCO_3_ (1 mL). The layers were separated and the organic layer was washed with brine (1 mL), dried over MgSO_4_ and concentrated under reduced pressure. Without further purification the residue was dissolved in dry MeCN (15 mL), **10** (530 mg, 3.02 mmol) was added and the mixture was cooled to -30 ^o^C. Sn(OTf)_2_ (50.4 mg, 0.12 mmol) was added and the mixture was allowed to warm up to RT overnight. The mixture was dilute with EtOAC (50mL) and quenched with sat. aq. NaHCO_3_ (1 mL). The layers were separated and the organic layer was dried over MgSO_4_ and concentrated under reduced pressure. The resulting residue was purified by column chromatography (EtOAc:heptane = 1:3) to yield 392 mg of a white solid of **13** (0.95 mmol, 79%) with recovery of 41 mg starting material **12** (0.12 mmol, 10 %). *R*_F_ = 0.49 (1:1, EtOAc:heptane); ^1^H NMR (500 MHz, CDCl3) δ 7.35 (q, *J* = 8.9, 7.6 Hz, 10H), 5.33 – 5.04 (m, 4H), 4.91 (m, 1H), 4.68 (dp, *J* = 11.2, 3.9 Hz, 1H), 2.99 – 2.40 (m, 2H), 2.30 (s, 1H), 2.24 – 1.75 (m, 3H), 1.76 – 1.61 (m, 3H), 1.57 – 1.50 (m, 1H), 1.47 – 1.24 (m, 1H); ^13^C NMR (126 MHz, CDCl_3_) δ 172.7, 136.6, 128.8, 128.6, 128.5, 128.2, 128.0, 67.6, 67.2, 53.5, 46.7, 45.8, 30.4, 30.2, 26.3, 15.7; HRMS (ESI+): calcd for C_22_^13^C_2_H_27_NNaO_5_ (*M*+Na^+^): 434.1854, found 434.1857.

**(2*S*)-6-(2-Oxopropyl-1,3-^13^C_2_)piperidine-2-carboxylic acid [14]**

To a degassed solution of **9** (382 mg, 0.90 mmol) in MeOH:water (7 ml, 6:1) was added Pd/C (15 mg, 0.14 mml, 10 mol% Pd). The atmosphere was exchanged to hydrogen and the suspension was stirred for 4 h. The reaction mixture was filtered over Celite, concentrated under reduced pressure. The product was purified by chromatography on Iatro beads (0 -> 40% MeOH in DCM) and lyophilized to obtain 128 mg of **14** (0.68 mmol, 76%) as a white powder. ^1^H NMR (500 MHz, D_2_O) δ 3.66 (dd, *J* = 12.4, 3.3 Hz, 1H), 3.60 (ddq, *J* = 12.5, 6.3, 3.2 Hz, 1H), 3.24 – 2.80 (m, 2H), 2.33 – 2.23 (m, 1H), 2.27 (dd, ^1^*J_CH_* = 128.3, ^3^*J* = 1.7 Hz, 3H), 2.05 – 1.86 (m, 2H), 1.73 – 1.40 (m, 3H); ^13^C NMR (126 MHz, deuterium oxide) δ 60.2, 45.6, 45.5, 41.2, 29.7, 29.6, 27.7, 26.5, 26.3, 22.9, 22.1, 21.8, 21.5; HRMS (ESI+): calcd for C_7_^13^C_2_H_16_NO_3_(*M*+H^+^): 188.1197, found 188.1198.
